# Genetically downregulated interleukin-6 signaling is associated with a favorable cardiometabolic profile: a phenome-wide association study

**DOI:** 10.1101/2020.10.28.20220822

**Authors:** Marios K. Georgakis, Rainer Malik, Xue Li, Dipender Gill, Michael G. Levin, Ha My T. Vy, Renae Judy, Marylyn Ritchie, Shefali S. Verma, Regeneron Genetics Center, Girish N. Nadkarni, Scott M. Damrauer, Evropi Τheodoratou, Martin Dichgans

**Affiliations:** Institute for Stroke and Dementia Research (ISD), University Hospital, Ludwig-Maximilians-University LMU, Munich, Germany; Centre of Global Health, Usher Institute, University of Edinburgh, Edinburgh, UK; School of Public Health and the Second Affiliated Hospital, Zhejiang University, Hangzhou, China; Department of Epidemiology and Biostatistics, School of Public Health, Imperial College London, London, United Kingdom; Division of Cardiovascular Medicine, Department of Medicine, Perelman School of Medicine, University of Pennsylvania, Philadelphia, USA; The Charles Bronfman Institute for Personalized Medicine, Icahn School of Medicine at Mount Sinai, New York, USA; Department of Surgery, Perelman School of Medicine, University of Pennsylvania, Philadelphia, USA; Department of Genetics, Perelman School of Medicine, University of Pennsylvania, Philadelphia, USA; Regeneron Genetics Center, Tarrytown, New York, USA; Hasso Plattner Institute for Digital Health at Mount Sinai, Icahn School of Medicine at Mount Sinai, New York, USA; Department of Medicine, Icahn School of Medicine at Mount Sinai, New York, USA; Department of Surgery, Corporal Michael Crescenz VA Medical Center, Philadelphia, Pennsylvania, USA; Edinburgh Cancer Research Centre, Institute of Genetics and Molecular Medicine, University of Edinburgh, Edinburgh, UK; Munich Cluster for Systems Neurology (SyNergy), Munich, Germany; German Centre for Neurodegenerative Diseases (DZNE), Munich, Germany

**Keywords:** interleukin-6, inflammation, atherosclerosis, diabetes, cardiovascular disease, high-density lipoprotein cholesterol

## Abstract

**Background:** Interleukin-6 (IL6) signaling is a key inflammatory pathway widely implicated in the pathogenesis of multiple diseases including autoimmune, vascular, and metabolic disorders. While IL6-receptor (IL6R) inhibitors are already in use for the treatment of autoimmune diseases, their repurposing potential and safety profile is still debated.

**Methods:** We used 7 genetic variants at the *IL6R* locus as proxies for IL6 signaling downregulation and explored their effects on 1,428 clinical outcomes in a phenome-wide association study (PheWAS) using data from the UK Biobank (339,256 unrelated individuals). Significant associations were meta-analyzed with data from the Penn Medicine (10,244 individuals) and BioMe (9,054 individuals) Biobanks for validation. We further investigated associations between genetically downregulated IL6 signaling and 366 biomarkers and endophenotypes of human disease in the UK Biobank and other phenotype-specific consortia. All associations were examined by Mendelian randomization (MR) analyses scaled to the effects of tocilizumab, a monoclonal antibody targeting IL6R.

**Results:** The PheWAS-MR analyses showed significant associations with 16 clinical outcomes and 17 biomarkers following correction for multiple comparisons. Genetically downregulated IL6 signaling was associated with a lower risk of several atherosclerotic phenotypes including ischemic heart disease (OR: 0.84, 95%CI: 0.77-0.90) and abdominal aortic aneurysm (OR: 0.44, 95%CI: 0.29-0.67). We further found significant associations with lower risk of type 2 diabetes (OR: 0.80, 95%CI: 0.73-0.88), lower glycated hemoglobin A1c (HbA1c) levels (beta: −0.07, 95%CI: −0.08 to −0.05), and higher HDL-cholesterol levels (beta: 0.04, 95%CI: 0.02-0.06). In accord with clinical trials examining pharmacological IL6 blockade, genetically downregulated IL6 signaling was associated with higher risk of neutropenia and bacterial infections (cellulitis and urinary tract infections) and with higher hemoglobin concentrations. We further found significant associations with higher risk of atopic dermatitis and higher levels of the pro-allergic cytokine interleukin-4.

**Conclusions:** Genetic IL6 signaling downregulation associates with a lower risk of vascular outcomes and a more favorable cardiometabolic profile. These findings further support a repurposing of IL6R blockade for lowering cardiovascular risk while also informing on potential side effects.

## Introduction

Interleukin-6 (IL6) is a key cytokine that is involved in the activation and regulation of immune responses, tissue regeneration, and metabolism.^1^ IL6 plays a critical role in the pathogenesis of multiple inflammatory disorders^2, 3^ and anti-inflammatory approaches targeting the IL6 signaling cascade have gained great attention. Antibodies against the IL6 receptor (IL6R) are used for the treatment of rheumatoid arthritis^4^ and other autoimmune disorders.^5, 6^ Accumulating experimental, epidemiological, and clinical evidence further supports a broader role of IL6 signaling in human disease, as has specifically been demonstrated for cardiovascular, metabolic, and psychiatric disorders.^3, 7, 8^

Still, it remains unknown whether pharmacological IL6R blockade could be effectively repurposed for the treatment or prevention of diseases beyond current indications. Also, any potential benefits would need to be balanced against the side-effects of such approaches. Both tocilizumab and sarilumab, the IL6R inhibitors currently in use, have been found to be associated with an increased rate of infections,^9^ hyperlipidemia^4, 10^ and hepatotoxicity.^4, 9^ Yet, it remains unknown whether these adverse effects are inherently linked to the downregulation of IL6 signaling or represent compound-specific properties. Furthermore, less common side-effects could have remained undetected in previous clinical trials.

A comprehensive understanding of the repurposing opportunities and unknown side-effects of downregulation of IL6 signaling would help prioritizing indications that should be moved forward to clinical trials. While observational studies could provide such information, the risk of bias due to confounding by indication remains high in observational settings.^11^ The growing availability of genome-wide association studies (GWAS) and the development of large-scale biobanks linked to health records offer the possibility to study drug effects by use of genetic data.^12-14^ Specifically, Mendelian randomization (MR) allows the use of genetic variants to proxy traits of interest and explore widespread associations with outcomes in phenome-wide association studies (PheWAS).^15^ MR overcomes key limitations of observational studies, such as confounding and reverse causation, and allows for investigation of causal effects on outcomes.^16, 17^

We recently identified a set of 7 genetic variants in the locus of the *IL6R* gene that showed remarkably similar effects on upstream (soluble IL6R and IL6) and downstream (C-reactive protein [CRP] and fibrinogen) molecules in the IL6 signaling cascade compared to those derived from clinical trials for tocilizumab.^18^ Here, we used these variants as a proxy of IL6 signaling downregulation and performed an MR-PheWAS study to explore potential repurposing opportunities and unknown side-effects associated with IL6R blockade. We first examined effects on clinical outcomes in a sample of 339,256 White British individuals from the population-based UK Biobank study. We subsequently examined the identified signals in two independent samples of 10,244 and 9,054 European American individuals from the Penn Medicine Biobank (PMBB) and the BioMe Biobank in the US, respectively. Finally, we explored the effects of genetically downregulated IL6 signaling on multiple disease-related biomarkers including hematological, biochemical, metabolomic, inflammatory, immunological, hemodynamic, and anthropometric traits in data from phenotype-specific genetic consortia.

## Methods

### Data sources and Ethics statement

This study follows the MR-STROBE guidelines.^19^ Data for clinical outcomes are from UK Biobank. Data from Penn Medicine and BioMe Biobanks were used for validation. Data for biomarkers were obtained from the UK Biobank or publicly available summary statistics. Supporting data for the UK Biobank analyses are available in the online-only Data Supplement. Supporting data from Penn Medicine and BioMe Biobanks are available upon request. Analyses were performed in R (v3.5.0; The R Foundation for Statistical Computing).

Participants in all studies provided informed consent and all studies obtained ethical approval from institutional review boards. We accessed individual-level data from the UK Biobank data following approval of an application by the UK Biobank Ethics and Governance Council (application #2532). For the Penn Medicine and BioMe Biobanks, individual-level data were analyzed from investigators of the studies (ML, RJ, MR, SV, SD for Penn Medicine and HMV, GN for BioMe) after receiving approval for the current study from the University of Pennsylvania Institutional Review Board and the Institutional Review Board of the Icahn School of Medicine, respectively. We then accessed the derived summary statistics to meta-analyze them with the UK Biobank data.

### Selection of genetic instruments

The exposure trait in this study was IL6R-mediated downregulation of the IL6 signaling cascade. We used as genetic instruments 7 single nucleotide polymorphisms (SNPs) within or close to the *IL6R* gene, which have previously been validated to proxy the biochemical effects of pharmacological IL6R inhibition.^18^ Instrument selection is detailed elsewhere.^18^ Briefly, variants were located within the *IL6R* gene or a region 300 kB upstream or downstream of it (**Supplementary Figure 1**) and were selected on the basis of their associations with lower levels of CRP that is downstream to IL6R-mediated signaling and reflects the activity of the IL6 signaling pathway.^3, 20^ The variants were derived from a GWAS of 204,402 individuals of European ancestry,^21^ associated with circulating CRP levels at genome-wide significance (p<5×10^−8^) and were clumped for linkage disequilibrium at *r*^*2*^<0.1 according to the 1000G European reference panel. Aside from their effects on CRP levels the 7 genetic variants were also associated with higher levels of soluble IL6 receptor (sIL6R) and IL6, which are upstream molecules in the pathway, as well as with lower levels of fibrinogen, another downstream molecule in the IL6 signaling cascade.^18^ The effects of all 7 genetic variants on circulating sIL6R, IL6, CRP, and fibrinogen levels were directionally consistent with downregulation of the IL6 signaling pathway, as depicted in **Supplementary Figure 2**. Three of the selected variants were within the *IL6R* gene; to minimize emergence of potential pleiotropic effects through neighboring genes we performed sensitivity analyses restricted to these variants. All SNPs have been shown to be of sufficient strength (F>10) for use in MR analyses, thus minimizing risk of weak instrument bias.^18^ The genetic instruments are provided in **Supplementary Table 1** along with their effects on circulating CRP, fibrinogen, sIL6R, and IL6 levels. The proportion of variance of the circulating biomarker levels explained by each of the SNPs was estimated according to a previously described formula.^22^ The 7 SNPs explained 0.55% and 0.24% of the variance of CRP and fibrinogen levels, respectively, and 71.5% and 0.51% of the variance of sIL6R and IL6, respectively.

**Table 1.**
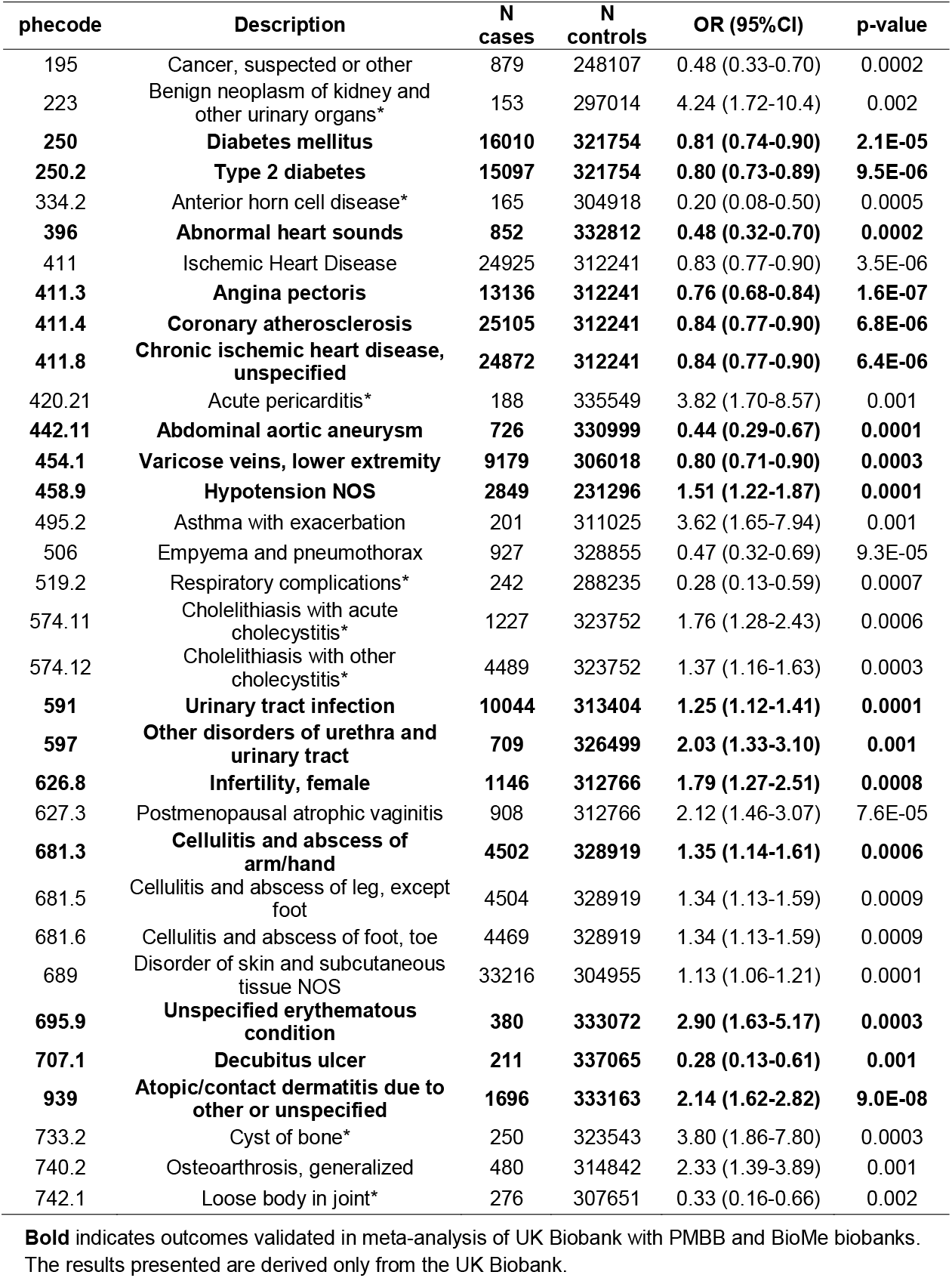
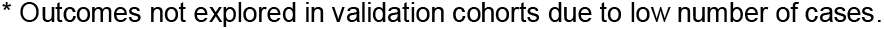
Mendelian Randomization results for clinical outcomes that were significantly associated with genetically downregulated IL6 signaling in UK Biobank. The Odds Ratios (OR) and 95% confidence intervals (95%CI) are derived from inverse-variance weighted Mendelian randomization and are scaled to the CRP-decreasing effect of tocilizumab.

| phecode | Description | N cases | N controls | OR (95%CI) | p-value |
| --- | --- | --- | --- | --- | --- |
| 195 | Cancer, suspected or other | 879 | 248107 | 0.48 (0.33-0.70) | 0.0002 |
| 223 | Benign neoplasm of kidney and other urinary organs* | 153 | 297014 | 4.24 (1.72-10.4) | 0.002 |
| **250** | **Diabetes mellitus** | **16010** | **321754** | **0.81 (0.74-0.90)** | **2.1E-05** |
| **250.2** | **Type 2 diabetes** | **15097** | **321754** | **0.80 (0.73-0.89)** | **9.5E-06** |
| 334.2 | Anterior horn cell disease* | 165 | 304918 | 0.20 (0.08-0.50) | 0.0005 |
| **396** | **Abnormal heart sounds** | **852** | **332812** | **0.48 (0.32-0.70)** | **0.0002** |
| 411 | Ischemic Heart Disease | 24925 | 312241 | 0.83 (0.77-0.90) | 3.5E-06 |
| **411.3** | **Angina pectoris** | **13136** | **312241** | **0.76 (0.68-0.84)** | **1.6E-07** |
| **411.4** | **Coronary atherosclerosis** | **25105** | **312241** | **0.84 (0.77-0.90)** | **6.8E-06** |
| **411.8** | **Chronic ischemic heart disease, unspecified** | **24872** | **312241** | **0.84 (0.77-0.90)** | **6.4E-06** |
| 420.21 | Acute pericarditis* | 188 | 335549 | 3.82 (1.70-8.57) | 0.001 |
| **442.11** | **Abdominal aortic aneurysm** | **726** | **330999** | **0.44 (0.29-0.67)** | **0.0001** |
| **454.1** | **Varicose veins, lower extremity** | **9179** | **306018** | **0.80 (0.71-0.90)** | **0.0003** |
| **458.9** | **Hypotension NOS** | **2849** | **231296** | **1.51 (1.22-1.87)** | **0.0001** |
| 495.2 | Asthma with exacerbation | 201 | 311025 | 3.62 (1.65-7.94) | 0.001 |
| 506 | Empyema and pneumothorax | 927 | 328855 | 0.47 (0.32-0.69) | 9.3E-05 |
| 519.2 | Respiratory complications* | 242 | 288235 | 0.28 (0.13-0.59) | 0.0007 |
| 574.11 | Cholelithiasis with acute cholecystitis* | 1227 | 323752 | 1.76 (1.28-2.43) | 0.0006 |
| 574.12 | Cholelithiasis with other cholecystitis* | 4489 | 323752 | 1.37 (1.16-1.63) | 0.0003 |
| **591** | **Urinary tract infection** | **10044** | **313404** | **1.25 (1.12-1.41)** | **0.0001** |
| **597** | **Other disorders of urethra and urinary tract** | **709** | **326499** | **2.03 (1.33-3.10)** | **0.001** |
| **626.8** | **Infertility, female** | **1146** | **312766** | **1.79 (1.27-2.51)** | **0.0008** |
| 627.3 | Postmenopausal atrophic vaginitis | 908 | 312766 | 2.12 (1.46-3.07) | 7.6E-05 |
| **681.3** | **Cellulitis and abscess of arm/hand** | **4502** | **328919** | **1.35 (1.14-1.61)** | **0.0006** |
| 681.5 | Cellulitis and abscess of leg, except foot | 4504 | 328919 | 1.34 (1.13-1.59) | 0.0009 |
| 681.6 | Cellulitis and abscess of foot, toe | 4469 | 328919 | 1.34 (1.13-1.59) | 0.0009 |
| 689 | Disorder of skin and subcutaneous tissue NOS | 33216 | 304955 | 1.13 (1.06-1.21) | 0.0001 |
| **695.9** | **Unspecified erythematous condition** | **380** | **333072** | **2.90 (1.63-5.17)** | **0.0003** |
| **707.1** | **Decubitus ulcer** | **211** | **337065** | **0.28 (0.13-0.61)** | **0.001** |
| **939** | **Atopic/contact dermatitis due to other or unspecified** | **1696** | **333163** | **2.14 (1.62-2.82)** | **9.0E-08** |
| 733.2 | Cyst of bone* | 250 | 323543 | 3.80 (1.86-7.80) | 0.0003 |
| 740.2 | Osteoarthritis, generalized | 480 | 314842 | 2.33 (1.39-3.89) | 0.001 |
| 742.1 | Loose body in joint* | 276 | 307651 | 0.33 (0.16-0.66) | 0.002 |

6 **Bold** indicates outcomes validated in meta-analysis of UK Biobank with PMBB and BioMe biobanks. 7 The results presented are derived only from the UK Biobank.
\* Outcomes not explored in validation cohorts due to low number of cases.

**Figure 1.**
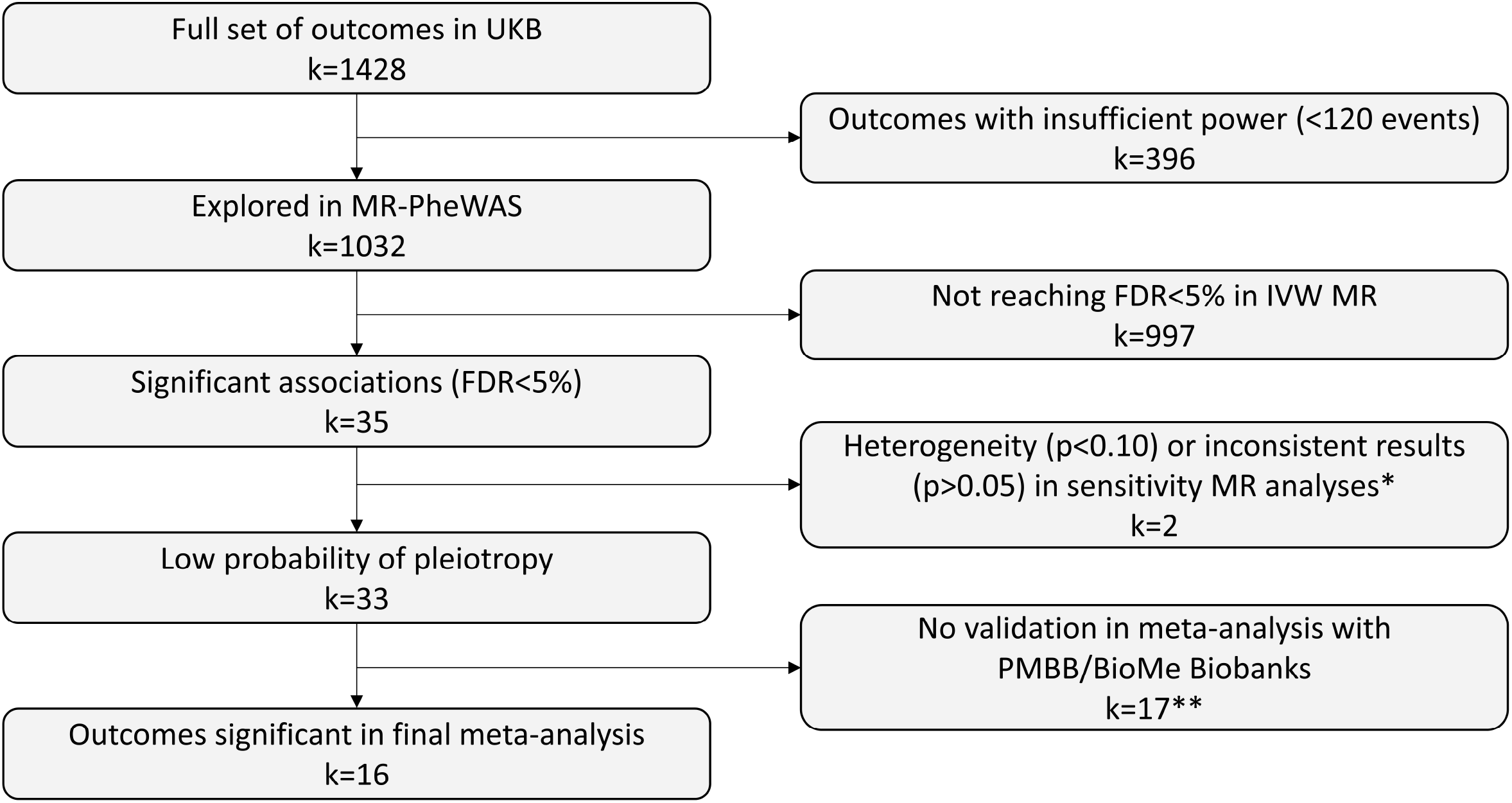
Flowchart of the significant associations for clinical outcomes. *k* refers to number of outcomes *weighted median, IVW restricted to 3 SNPs within IL6R **9 outcomes non-significant in meta-analysis & 8 outcomes not available for analysis in validation cohorts

**Figure 2.**
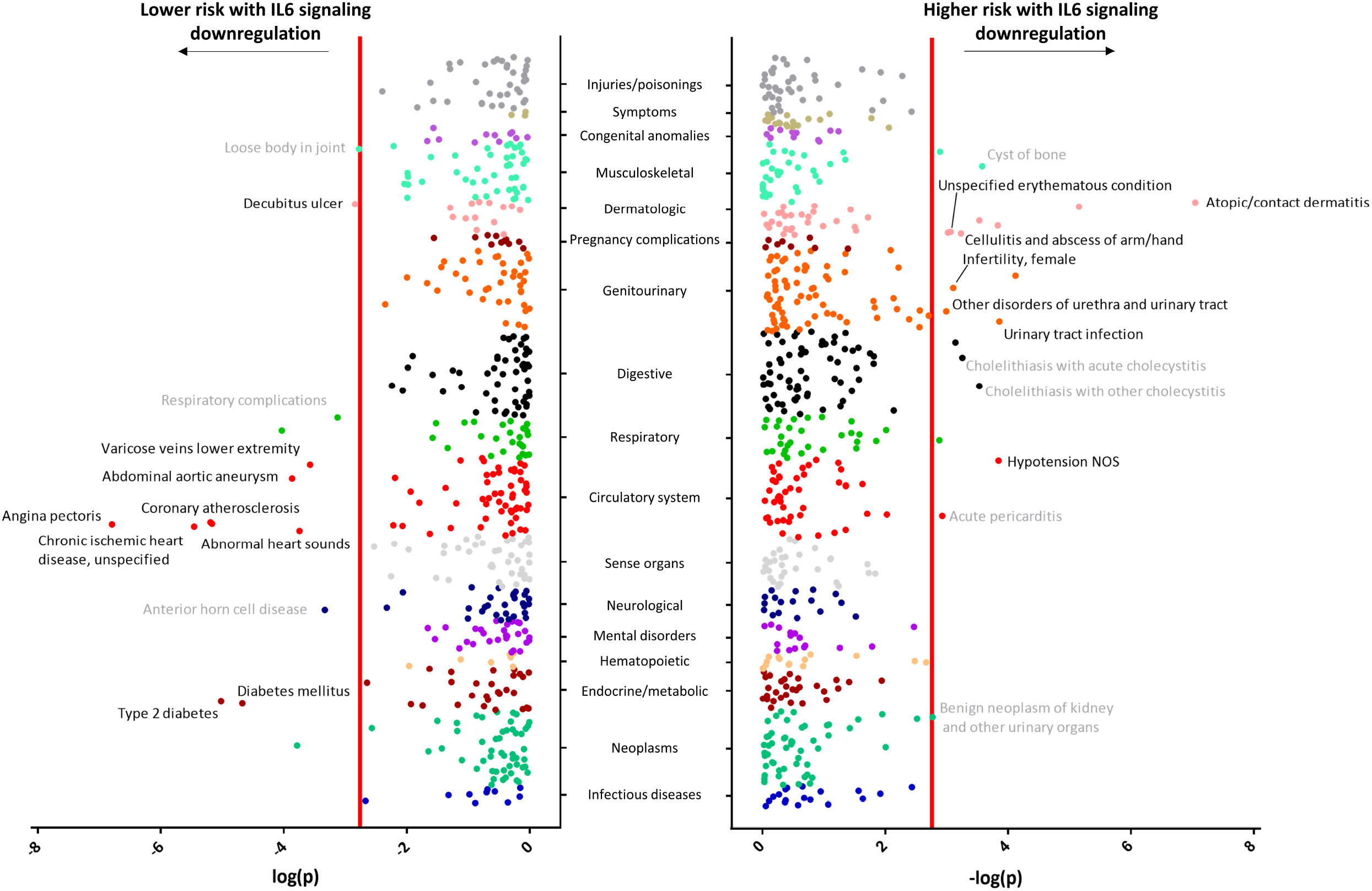
Results of the phenome-wide association study (PheWAS) for clinical outcomes in the UK Biobank and validation in PMBB and BioMe Biobanks. Shown are the results from the inverse-variance weighted Mendelian randomization analyses in the UK Biobank. The x-axes correspond to the logarithms of the p-values derived from these analyses. The red lines correspond to the statistical significance level (FDR<0.05). Outcomes surviving all significance criteria (Figure 1) in the UK Biobank analyses are labeled by name. Outcomes that were further validated in the meta-analysis of UK Biobank with PMBB and BioMe biobanks are labeled in black. Outcomes for which validation was unfeasible due to a low number of cases in both validation cohorts are labeled in grey.

### Phenome-wide association study in the UK Biobank

The UK Biobank is a population-based prospective cohort study that recruited 503,317 individuals aged 40-69 years in 2006-2010. Extensive baseline measurements including biochemical and genotype data were linked with national medical records (inpatient hospital episode records, primary general practitioner data, cancer registry, death registry) for longitudinal follow-up. The current analysis was constrained to a subset of unrelated White British individuals with high-quality genotype data in order to minimize the influence of diverse population structure within UK Biobank. The largest possible subset of individuals without relatedness was identified using an algorithm implemented in the R package “*i-graph (v1*.*0*.*1)*” developed by Bycroft and colleagues.^23^ UK Biobank genotype imputation was conducted based on a merged reference panel of the Haplotype Reference Consortium (HRC) panel, the UK10K panel and the 1000 Genome Phase 3 panel.^23^ All 7 SNPs used in current study were on the HRC panel. A total of 339,256 individuals were included in the PheWAS analyses.

Outcomes to be explored were defined by the PheCODE scheme.^24^ This scheme automatically excludes patients that have similar or potentially overlapping disease states from the corresponding control group. We used the International Classification of Diseases (ICD) versions 9 and 10 to identify cases in the medical records, with both incident and prevalent cases included. A map matching ICD-9 and −10 codes to phecodes was used, as previously described (https://phewascatalog.org/phecodes_icd10).^25^

We applied logistic regression to explore the effects of each SNP on each outcome adjusting for age, sex, the UK Biobank assessment center attended, east and north coordinates of home address, and the first five ancestral principal components. Based on power calculations, ≥120 cases would be required to detect odds ratios (OR) ≥2 or ≤0.5 at p<0.05 scaled to the CRP-decreasing effects of tocilizumab in the subsequent MR analyses (**Supplementary Figure 3**). Hence, all outcomes with <120 cases were excluded from analyses resulting in a total of 1,032 outcomes that were eventually included in the PheWAS analyses.

**Figure 3.**
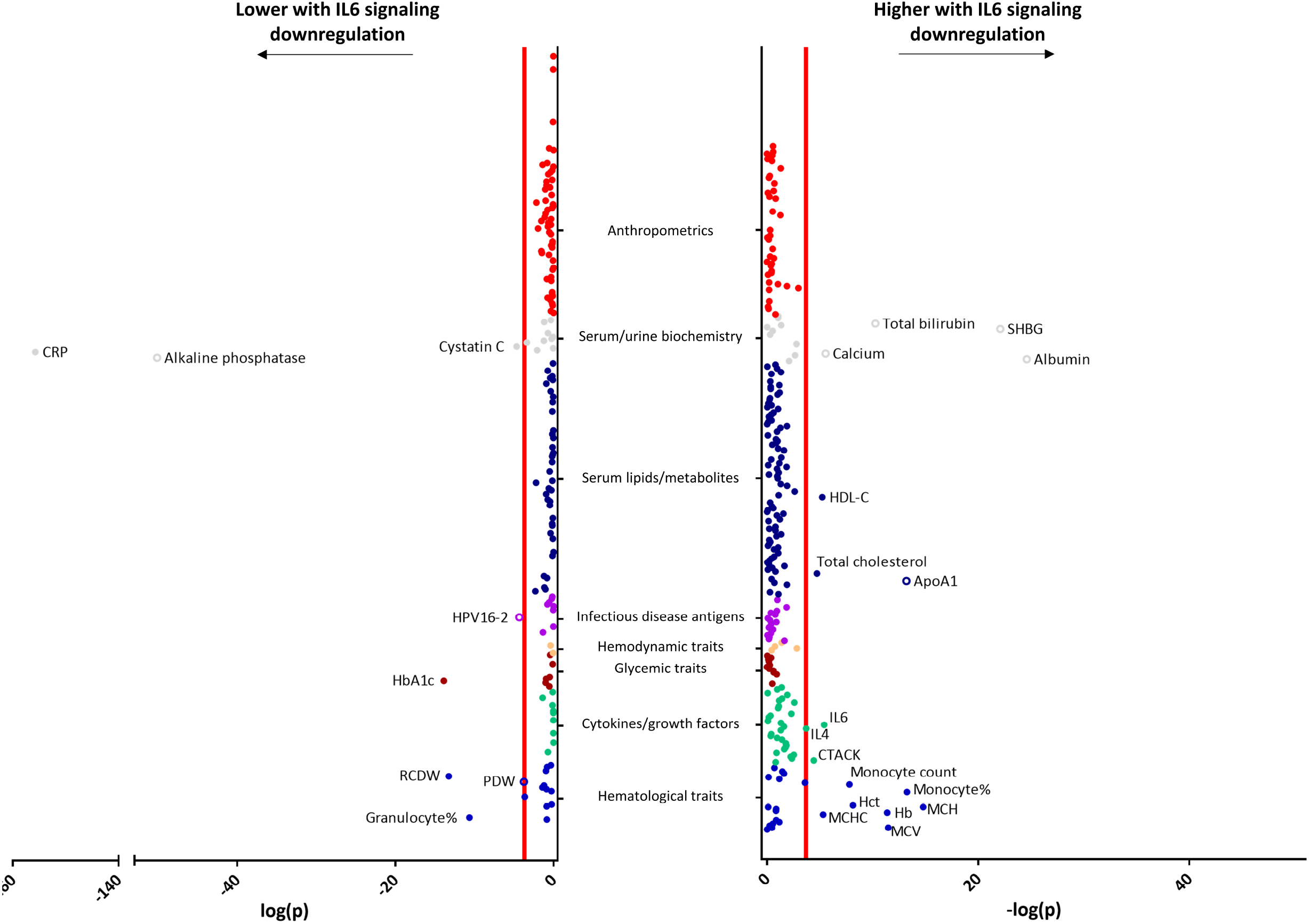
Results of the phenome-wide association study (PheWAS) for biomarkers. Shown are the results from inverse-variance weighted Mendelian randomization analyses. The x-axes correspond to the logarithms of the p-values derived from these analyses. The red lines correspond to the statistical significance level (FDR<0.05). Solid circles correspond to outcomes that survived all our significance criteria (**Supplementary Figure 2**). Open circles correspond to outcomes that showed significant associations while there was significant heterogeneity between the effects of the individual variants.

### Associations with biomarkers

We further derived association estimates of each SNP with the following biomarkers: (i) 25 serum and urine biomarkers available in the biochemistry panel of the UK Biobank (353,579 individuals); (ii) 36 hematological traits, as derived from the summary statistics of the study by Astle *et al*.^26^ based on the UK Biobank and the INTERVAL population (173,480 European individuals); (iii) 124 nuclear magnetic resonance (NMR)-measured serum lipids and metabolites, as derived from the publicly available summary statistics provided by Kettunen *et al*.^27^ (24,925 individuals of European ancestry); (iv) circulating levels of 41 cytokines and growth factors, derived from the publicly available summary statistics by Ahola-Olli *et al*.^28^ (8,293 individuals of Finnish ancestry); (v) 25 infectious disease antigens available in a sub-sample of 9,431 individuals from the UK Biobank; (vi) 6 hemodynamic traits that were available in the UK Biobank (408,228 individuals); (vii) 18 glycemic traits made publicly available from a series of analyses from the MAGIC Consortium (up to 133,010 individuals)^29^; and (viii) 91 anthropometric traits derived from the summary statistics made publicly available by the GIANT Consortium (different sample sizes, up to 322,154 individuals).^30^ The data sources for these studies are described in **Supplementary Table 2**.

**Table 2.** Mendelian Randomization results for biomarkers that were significantly associated with genetically downregulated IL6 signaling. The betas and 95% confidence intervals (95%CI) are derived from inverse-variance weighted Mendelian randomization and are scaled to the CRP-decreasing effect of tocilizumab.

| Description | beta (95%CI) | p-value |
| --- | --- | --- |
| C-reactive protein | -0.51 (-0.54 to -0.47) | 1.9E-156 |
| Cystatin C | -0.03 (-0.05 to -0.02) | 2.1E-05 |
| Granulocyte percentage of myeloid white cells | -0.10 (-0.13 to -0.07) | 2.2E-11 |
| Hematocrit | 0.08 (0.06 to 0.11) | 7.2E-09 |
| Hemoglobin concentration | 0.10 (0.07 to 0.13) | 4.2E-12 |
| Mean corpuscular hemoglobin | 0.12 (0.09 to 0.15) | 1.6E-15 |
| Mean corpuscular hemoglobin concentration | 0.07 (0.04 to 0.09) | 4.8E-06 |
| Mean corpuscular volume | 0.10 (0.08 to 0.13) | 3.3E-12 |
| Monocyte count | 0.08 (0.06 to 0.11) | 1.6E-08 |
| Monocyte percentage of white cells | 0.11 (0.08 to 0.14) | 5.6E-14 |
| Red cell distribution width | -0.11 (-0.14 to -0.08) | 5.6E-14 |
| Cutaneous T-cell-attracting chemokine | 0.41 (0.21 to 0.60) | 3.7E-05 |
| Interleukin-4 | 0.26 (0.13 to 0.39) | 0.0002 |
| Interleukin-6 | 0.31 (0.18 to 0.44) | 3.8E-06 |
| Glycated haemoglobin (HbA1c) | -0.07 (-0.08 to -0.05) | 1.3E-14 |
| HDL Cholesterol | 0.04 (0.02 to 0.06) | 5.9E-06 |
| Cholesterol | 0.04 (0.02 to 0.06) | 1.9E-05 |

For the genetic analyses of serum and urine biomarkers in the UK Biobank we performed linear regression, adjusting for age, sex, genotyping chip, assessment center and the first 20 ancestral principal components. Continuous traits were quantile-normalized, forcing phenotypes to a N(0, 1) distribution. For the infectious disease antigens, we performed logistic regression analyses specifying as cases all those individuals surpassing pre-specified cutoffs for seropositivity for single or multiple antigens as suggested by UK Biobank (available at https://biobank.ctsu.ox.ac.uk/crystal/crystal/docs/infdisease.pdf) using the same covariates as for the linear regression analysis. To ensure numerical stability, we performed linear transformation of covariates to a distribution of mean=0 and variance=1. The details of the GWAS analyses for the remaining traits that were derived from publicly available summary statistics are provided in the original publications (**Supplementary Table 2**).

### Statistical analyses – Mendelian randomization

We used the SNP-specific association estimates for the clinical outcomes and the biomarkers that we derived from the abovementioned analyses or that we extracted from the respective summary statistics and ran inverse-variance weighted (IVW) MR analyses to obtain a composite effect of IL6 signaling downregulation on the outcomes of interest.^31^ To provide clinically relevant results, the composite effect estimates were scaled to the CRP-decreasing effect of tocilizumab (a decrease of CRP levels by 67% on a dose of 8 mg/kg, 4 to 24 weeks post-administration).^20^ We corrected for multiple comparisons with the false discovery rate (FDR) method. We considered as statistically significant all those outcomes for which q-values were <0.05.

IVW MR was our primary approach. While the selection of instruments on a specific gene reduces the possibility of invalid variants,^32^ the derived estimates might still be biased in case of directional pleiotropy. Hence, for outcomes reaching statistical significance in the main analyses, we assessed heterogeneity across the SNP-specific MR estimates in the IVW MR analyses with the Cochran’s Q statistic as a measure of overall pleiotropy.^33^ For significant outcomes, we further applied sensitivity MR analyses that are more robust to the inclusion of pleiotropic variants. First, we used the weighted median estimator, which provides consistent estimates as long as at least half of the variants used in the MR analysis are valid.^34^ Second, we restricted our selection of instruments to SNPs within the *IL6R* gene, to avoid potential pleiotropic effects through genes neighboring *IL6R* and increase confidence in the effects of the instruments through IL6 signaling. As finally significant results, we considered all those that (i) showed an FDR-derived q<0.05 in the main IVW analyses, (ii) showed no heterogeneity (Cochran Q-derived p>0.10), and (iii) showed directionally consistent associations of p<0.05 in both sensitivity analyses.

### Validation in the Penn Medicine and BioMe Biobanks

For phenotypes reaching statistical significance after FDR control in original analyses, we further examined their associations with genetically downregulated IL6 signaling in the PMBB (University of Pennsylvania) and the BioMe Biobank (Icahn School of Medicine, Mount Sinai). PMBB is a longitudinal genomics and precision medicine study in which participants consent to linkage of genomic information and biospecimens to electronic health records; more than 60,000 individuals are currently enrolled. The BioMe Biobank is an electronic health record-linked clinical care biobank cohort that enrolls participants non-selectively from across the Mount Sinai Health System; the data set comprises over 45,000 participants from diverse ancestries (African, Hispanic/Latino, European and Other ancestries), with accompanying genome-wide genotyping data and a wide array of phenotypes derived from interview-based questionnaire, electronic health records and ICD-codes. For the current analyses, we considered only individuals of European ancestry, who underwent genome-wide genotyping and had available electronic health record data (PMBB: 10,244 and BioMe: 9,054). Clinical diagnoses in both biobanks were also coded according to the PheCODE schema. The MR analyses were repeated in these cohorts and the effect estimates from the IVW analyses were then meta-analyzed with those derived from the UK Biobank using random-effects models. Outcomes that reached the original FDR-corrected threshold in the meta-analysis of UKB, PMBB and BioMe were considered statistically significant.

## Results

### PheWAS: associations of IL6 signaling downregulation with clinical outcomes

The baseline descriptive characteristics of the UK Biobank participants that were included in the PheWAS analyses are presented in **Supplementary Table 3**. Baseline characteristics of individuals included in the validation cohorts (PMBB and BioMe biobanks) are presented in **Supplementary Table 4**. The number of outcomes examined per diagnostic category are presented in **Supplementary Table 5**, whereas the results of the MR analyses along with the number of cases and controls considered per outcome are provided in **Supplementary Table 6**. There were 35 outcomes reaching the statistical significance threshold of FDR<0.05 in the primary IVW MR analyses (p<1.7×10^−3^) that pooled the effect estimates across the effects of the 7 SNPs used as instruments (**Figure 1**). Out of those, 33 (94%) showed no evidence of heterogeneity (p>0.10) while exhibiting consistent associations (same direction and p<0.05) in the sensitivity MR analyses (weighted median, gene-restricted IVW), thus showing low probability of pleiotropy (**Supplementary Table 7** and **Figure 1**).

The detailed results from the inverse-variance weighted analyses for these outcomes are presented in **Table 1**. In the meta-analysis of the effect estimates derived from the UK Biobank with those from the PMBB and BioMe Biobanks, 16 of the 24 outcomes with sufficient statistical power for validation remained statistically significant (p<1.7×10^−3^; **Figure 2** and detailed results in **Supplementary Table 8**). Most of the significant results (7/16, 44%) referred to outcomes related to cardiovascular disease. Among them, there were associations of genetically downregulated IL6 signaling with lower risk of angina pectoris, ischemic heart disease, coronary atherosclerosis, abdominal aortic aneurysm, abnormal heart sounds, and varicose veins of the lower extremities. Furthermore, we found associations with lower risk of diabetes mellitus, and particularly type 2 diabetes mellitus, as well as with lower risk of decubitus ulcer. On the opposite direction, we found associations of genetically downregulated IL6 signaling with higher risk of cellulitis and abscess of arm/hand, urinary tract infections, other disorders of urethra and urinary tract, female infertility, unspecified erythematous conditions, atopic dermatitis, and hypotension.

For nine additional outcomes that reached statistical significance in the primary UK Biobank analysis (benign neoplasm of kidney and other urinary organs, anterior horn cell disease, acute pericarditis, respiratory complications, cholelithiasis with acute cholecystitis or other cholecystitis, cyst of bone, loose body in joint), there were no sufficient number of cases in the PMBB and BioMe biobanks to allow for validation (**Table 1** and **Supplementary Table 8**).

### Associations with biomarkers

To gain additional insights into the relationship between IL6 signaling and subclinical endophenotypes relevant to human disease, we explored associations between genetically downregulated IL6 signaling and eight categories of 366 biomarkers. The results, along with the number of individuals examined in each analysis are presented in **Supplementary Table 9**. In the main IVW analysis, a total of 25 outcomes reached the threshold of statistical significance of FDR<0.05 (p<1.8×10^−4^). Of them, 17 associations remained consistent (same direction and p<0.05) in sensitivity analyses and did not show any heterogeneity (p>0.10) (**Supplementary Table 10** and **Supplementary Figure 4**).

**Table 2** displays the results of the IVW MR analyses for these biomarkers, which are also graphically presented in **Figure 3**. Aside from the expected associations with higher CRP and lower IL6 levels that we have previously reported,^18^ we found associations with a number of hematological traits. Specifically, genetically downregulated IL6 signaling was associated with higher mean corpuscular hemoglobin and volume, higher hemoglobin concentration, higher hematocrit, and lower red cell distribution width, as well as with higher monocyte count and percentage, and lower granulocyte percentage. Furthermore, we found genetically downregulated IL6 signaling to be associated with a lower glycated hemoglobin (HbA1c). Among serum lipids and metabolites, we found genetically downregulated IL6 signaling to be associated with a higher total cholesterol concentration and higher HDL cholesterol (**Supplementary Figure 5**). There were also significant associations with lower cystatin C, and with higher levels of the cytokines IL4 (interleukin-4) and CTACK (Cutaneous T-Cell Attracting Chemokine).

## Discussion

This study used large-scale data from multiple sources to explore the phenotypic consequences of genetic predisposition to IL6 signaling downregulation. Our findings can be summarized as follows: genetic downregulation of IL6 signaling was associated with (i) lower risk of atherosclerotic vascular phenotypes (coronary artery disease and abdominal aortic aneurysm), (ii) lower HbA1c and lower risk of type 2 diabetes mellitus, (iii) increases in cholesterol levels, mainly due to an effect on HDL cholesterol, (iv) higher risk of skin and urinary tract infections and neutropenia, (v) higher risk of atopic phenotypes and higher interleukin-4 levels, and (vi) increases in hemoglobin and related phenotypes, as well as monocyte counts.

As an important novel finding, we provide evidence for a causal effect of IL6 signaling on the risk of type 2 diabetes. Aside from effects on risk of type 2 diabetes in three independent cohorts, we also found directionally consistent effects on HbA1c levels, thus further validating our results. Our findings complement and expand existing data from other sources supporting an involvement of IL6 signaling in the development of hyperglycemia and diabetes. Analyses of case series of diabetic and non-diabetic patients with rheumatoid arthritis have shown that pharmacological IL6R inhibition leads to reductions in HbA1c levels.^35, 36^ Moreover, meta-analyses of cohort studies show significant associations between elevated IL6 levels and incident type 2 diabetes.^37^ Experimental studies in mice further suggest an effect of IL6 on insulin resistance that can be reversed by IL6 signaling blockade.^38^ As such, there is triangulation of evidence to support IL6-centered approaches as a strategy for lowering risk of hyperglycemia and type 2 diabetes. Future trials on IL6R inhibition should add glycemic traits and incident type 2 diabetes to the outcomes under study.

Clinical trials exploring pharmacological IL6R blockade have reported increases in total cholesterol levels.^4, 10^ However, it remains unclear how IL6R blockade influences the different lipid fractions, which have been shown to differentially influence vascular risk.^39-41^ Here, we explored associations with a panel of >100 lipid fractions and metabolites. Although we found genetic predisposition to downregulation of IL6 signaling to be indeed associated with higher total cholesterol levels, this increase was largely restricted to HDL- and not LDL-cholesterol. We further found a significant association with Apolipoprotein-A1, the hallmark of HDL particles, but not Apolipoprotein-B that is contained in non-HDL particles (**Supplementary Figure 5**). While clinical trials have shown no benefit of HDL-raising approaches for large vessel disease,^42^ recent evidence suggests that genetic elevation of HDL cholesterol levels is associated with lower risk of cerebral small vessel disease^40^ and chronic kidney disease.^43^ Whether these associations could be clinically meaningful remains to be tested in future clinical trials. Still, the association of IL6 signaling downregulation with lower risk of type 2 diabetes and higher HDL levels, both key components of the current definition of metabolic syndrome, suggest a beneficial effect of IL6R blockade on the metabolic profile of individuals at risk for cardiovascular disease.

The associations between genetically downregulated IL6 signaling and lower risk of multiple phenotypes related to coronary artery disease (coronary atherosclerosis, angina pectoris, chronic ischemic heart disease) and aortic aneurysm agrees with previous clinical,^44^ genetic,^18, 20, 45^ and experimental^46, 47^ data demonstrating a role of IL6 signaling in atherosclerosis. Accumulating data from clinical trials also support the efficacy of targeting specific inflammatory pathways for lowering vascular risk.^48, 49^ The CANTOS trial studied a monoclonal antibody targeting IL1β, a cytokine upstream to IL6, and demonstrated reductions in the rates of vascular events among patients with a history of myocardial infarction.^48^ Secondary analyses from CANTOS showed that the residual vascular risk after IL1β inhibition could be explained by IL6 levels,^44^ thus supporting direct IL6 signaling inhibition as a potentially even more effective strategy.^7^ By showing that genetic IL6 signaling downregulation associates with both a more favorable cardiometabolic profile and a decreased risk of vascular endpoints, our findings provide additional support towards repurposing IL6R blockade approaches for lowering vascular risk.

Aside from demonstrating cardiometabolic effects of IL6 signaling, our MR approach captured some of the known side-effects of IL6R blockade that have previously been reported by clinical trials. Specifically, we found associations of genetic predisposition to IL6 signaling downregulation with higher risks of infections (cellulitis and urinary tract infections) and atopic dermatitis.^4, 9, 50^ By analysing biomarker data, we identified associations with specific endophenotypes that might mediate these findings. Specifically, genetically downregulated IL6 signaling was associated with neutropenia that might explain its effects on bacterial infections, but also with higher IL4 levels, a key cytokine in allergic reactions. The higher risk associated with atopic reactions was further highlighted by a significant association with asthma in the UK Biobank, which we could not validate in the independent cohorts. Given the high prevalence of atopic dermatitis and asthma in the general population,^51^ future research should explore in more detail the role of IL6 in atopic reactions and whether IL6R blockade could lead to exacerbations in patients suffering from these diseases.We further found genetically downregulated IL6 signaling to be associated with higher hemoglobin concentrations and a number of other related biomarkers. Similar effects have been previously reported for pharmacological IL6R blockade and have been attributed to improvements in chronic disease anemia associated with inflammation and autoimmune diseases.^52^ Additionally, genetic IL6 signaling downregulation was associated with monocyte count and percentage. This is a finding not previously reported and the clinical significance, as well as the underlying mechanisms need to be further evaluated.

An important methodological consideration of Mendelian randomization analyses includes the potentially pleiotropic effects of the genetic variants used in the analyses on clinical outcomes through pathways other than the IL6 signaling cascade. To address this, we restricted our analyses to the *IL6R* locus, thus minimizing effects through other pathways. Furthermore, we addressed this issue in sensitivity analyses by further restricting our selection to SNPs *within* the *IL6R* gene. If the 7 SNPs had effects on clinical outcomes that act through different pathways, it would be expected that these effects would not be similar across the 7 SNPs, and there would be heterogeneity in the pooled effect sizes. Hence, heterogeneity was a criterion for selecting “significant” outcomes. All outcomes showing significant heterogeneity in the analyses (p<0.10) were filtered out of significant results. We believe that this set of sensitivity analyses minimized the possibility of unrecognized pleiotropic effects.

Our study has several strengths. Utilizing the most recent genetic data and a recently published and significantly expanded set of instruments^18^ we were sufficiently powered to expand the results derived from previous efforts exploring the effects of genetically downregulated IL6 signaling on clinical outcomes.^20, 45^ By applying a pre-specified approach for filtering the significant outcomes in the current study including strict thresholds for statistical significance, consistency across alternative statistical methods, lack of heterogeneity between individual variants, and validation in independent cohorts, we ensured robustness of our findings. Using CRP levels as a proxy for downstream IL6 signaling further enabled us to scale the derived association estimates to the reported effects of tocilizumab, thus providing clinically meaningful estimates that might be comparable to those obtained from future trials. Finally, we expanded our analyses to biomarkers and endophenotypes, allowing cross-validation of the findings across multiple levels of phenotypes.

Our study also has limitations. First, to proxy IL6 signaling we used CRP levels, which increase in response to the activation of the IL6 cascade.^53^ However, IL6 signaling is a complex cascade with a (exerted through membrane-bound IL6R expressed in limited tissues) and a trans-signaling component (exerted through the more widely expressed soluble IL6R).^53^ The two sub-pathways exert unique actions,^53^ but disentangling them with genetic instruments goes beyond the limits of MR. Second, by design, MR analyses assess the effects of lifetime downregulated IL6 signaling, which might differ from a shorter pharmacological inhibition with IL6R blockade. Third, there might be unknown pleiotropic effects of the genetic proxies used as instruments in the current study that might bias the reported associations. Of note, however, the results were remarkably consistent in sensitivity MR analyses that are more robust to the inclusion of pleiotropic variants, as well as in analyses restricted within the *IL6R* gene. Fourth, while the sample size of the discovery UK Biobank cohort was very large (>300,000 individuals) and provided power to explore associations with outcomes, the sample sizes of the validation cohorts were rather small and did not offer sufficient power to explore the robustness of nine signals for relatively rare outcomes that came up in the discovery cohort. Fifth, our results were solely based on individuals of European origin, and might thus not apply to other ethnicities. Sixth, while we used the largest available datasets to detect genetic variants that influence the IL6 signaling cascade, the proportion of the variance explained in biomarkers of the pathway was relatively low. As such, we cannot rule out that additional variants might influence the pathway and future analyses in larger datasets might provide additional insights towards detecting more such variants. Noteworthy, as long as the results are robust and not influenced by pleiotropy, we would not expect the number of genetic instruments used to influence the validity of the derived effects estimates. Finally, while genetic variants used as proxies for IL6 signaling influence CRP and other upstream and downstream molecules of the IL6 signaling cascade, the exact cellular and molecular significance of each variant remains unknown.

In conclusion, this study used large-scale data to systematically explore the phenotypic consequences of genetically downregulated IL6 signaling. We highlight key associations with a range of cardiometabolic traits including lower risks of type 2 diabetes and hyperglycaemia, higher HDL cholesterol levels, and lower risk of atherosclerotic outcomes. As such, our findings highlight the potential of repurposing IL6R blockade as a strategy for improving cardiometabolic profile and lowering vascular risk. These effects should be further explored in clinical trials and weighted against the side-effects of IL6-targeting approaches.

## Data Availability

The genetic variants used as instruments in the current study, as well as summary statistics derived for the examined outcomes are available as Supplementary Material. Raw UK Biobank data may be derived through application. Data from the Penn Medicine and BioMe Biobanks are also available upon application to principal investigators.

## Acknowledgements

This research has been conducted using the UK Biobank Resource (UK Biobank application 2532, “UK Biobank stroke study: developing an in-depth understanding of the determinants of stroke and its subtypes”). We thank the following studies and consortia for making summary data from GWASs publicly available: the INTERVAL study, the YFS and FINRISK studies, the MAGIC Consortium, the GWAS for NMR-measured metabolites by Kettunen *et al*., and the GIANT Consortium. We thank the personnel and the participants of the Penn Medicine and BioMe biobanks.

## Regeneron Genetics Center Banner Author List and Contribution Statements

All authors/contributors are listed in alphabetical order. RGC Management and Leadership

### Team

Goncalo Abecasis, Ph.D., Aris Baras, M.D., Michael Cantor, M.D., Giovanni Coppola, M.D., Aris Economides, Ph.D., John D. Overton, Ph.D., Jeffrey G. Reid, Ph.D., Alan R. Shuldiner, M.D. Sequencing and Lab Operations: Christina Beechert, Caitlin Forsythe, M.S., Erin D. Fuller, Zhenhua Gu, M.S., Michael Lattari, Alexander Lopez, M.S., John D. Overton, Ph.D., Thomas D. Schleicher, M.S., Maria Sotiropoulos Padilla, M.S., Karina Toledo, Louis Widom, Sarah E. Wolf, M.S., Manasi Pradhan, M.S., Kia Manoochehri, Ricardo H. Ulloa. Genome Informatics: Xiaodong Bai, Ph.D., Suganthi Balasubramanian, Ph.D., Leland Barnard, Ph.D., Andrew Blumenfeld, Yating Chai, Ph.D., Gisu Eom, Lukas Habegger, Ph.D., Young Hahn, Alicia Hawes, B.S., Shareef Khalid, Jeffrey G. Reid, Ph.D., Evan K. Maxwell, Ph.D., John Penn, M.S., Jeffrey C. Staples, Ph.D., Ashish Yadav, M.S. Planning, Strategy, and Operations: Paloma M. Guzzardo, Ph.D., Marcus B. Jones, Ph.D., Lyndon J. Mitnaul, Ph.D.

