## Supplementary Material for "Genetically downregulated interleukin-6 signaling is associated with a favorable cardiometabolic profile: a phenome-wide association study"

**Running title:** *Georgakis et al; Phenotypic outcomes of downregulated IL6 signaling*

Marios K. Georgakis, MD, PhD[1,2], Rainer Malik, PhD[1], Xue Li, PhD[3], Dipender Gill, BMBCh, PhD[4], Michael G. Levin, MD[5], Ha My T. Vy, PhD[6], Renae Judy, MS[7], Marylyn Ritchie, PhD[8], Shefali S. Verma, PhD[8], Regeneron Genetics Center[9], Girish N. Nadkarni, MD[6,10,11], Scott M. Damrauer, MD[7,12], Evropi Theodoratou, PhD[3,13], Martin Dichgans, MD[1,14,15]

[1]Institute for Stroke and Dementia Research (ISD), University Hospital, Ludwig-Maximilians-University LMU, Munich, Germany; [2]Graduate School for Systemic Neurosciences (GSN), Ludwig-Maximilians-University LMU, Munich, Germany; [3] Centre of Global Health, Usher Institute, University of Edinburgh, Edinburgh, UK; [4]Department of Epidemiology and Biostatistics, School of Public Health, Imperial College London, London, United Kingdom; [5]Division of Cardiovascular Medicine, Department of Medicine, Perelman School of Medicine, University of Pennsylvania, Philadelphia, USA; [6]The Charles Bronfman Institute for Personalized Medicine, Icahn School of Medicine at Mount Sinai, New York, USA; [7]Department of Surgery, Perelman School of Medicine, University of Pennsylvania, Philadelphia, USA; [8]Department of Genetics, Perelman School of Medicine, University of Pennsylvania, Philadelphia, USA; [9]Regeneron Genetics Center, Tarrytown, New York, USA; [10]Hasso Plattner Institute for Digital Health at Mount Sinai, Icahn School of Medicine at Mount Sinai, New York, USA; [11]Department of Medicine, Icahn School of Medicine at Mount Sinai, New York, USA; [12]Department of Surgery, Corporal Michael Crescenz VA Medical Center, Philadelphia, Pennsylvania, USA; [13]Edinburgh Cancer Research Centre, Institute of Genetics and Molecular Medicine, University of Edinburgh, Edinburgh, UK; [14]Munich Cluster for Systems Neurology (SyNergy), Munich, Germany; [14]German Centre for Neurodegenerative Diseases (DZNE), Munich, Germany

**Supplementary Table 1. Single nucleotide polymorphisms (SNP) used in the current analyses for proxying the effects of IL-6 signaling.** The betas, standard errors, and p-values refer to associations of these SNPs with CRP levels.

| | | | CRP | | | | | Fg | | | IL6 | | | sIl6R | | |
|---|---|---|---|---|---|---|---|---|---|---|---|---|---|---|---|---|
| **SNP** | **Effect allele** | **Non-effect allele** | **MAF** | **Beta †** | **SE** | **P** | **$R^2$ ‡** | **Beta †** | **SE** | **$R^2$ ‡** | **Beta †** | **SE** | **$R^2$ ‡** | **Beta †** | **SE** | **$R^2$ ‡** |
| rs73026617 | t | c | 0.097 | 0.0474 | 0.0068 | 3.2E-12 | 0.0003 | -0.0046 | 0.0015 | 7.3E-05 | 0.0135 | 0.0231 | 3.19E-05 | 0.2361 | 0.0416 | 0.0098 |
| rs12083537* | a | g | 0.193 | 0.0643 | 0.0053 | 7.1E-34 | 0.0009 | -0.0074 | 0.0012 | 0.0003 | 0.0053 | 0.0194 | 8.75E-06 | 0.025 | 0.0308 | 0.0002 |
| rs4556348* | t | c | 0.148 | 0.0541 | 0.0067 | 6.8E-16 | 0.0005 | -0.0053 | 0.0015 | 0.0001 | 0.0186 | 0.0214 | 8.72E-05 | 0.2 | 0.0367 | 0.0101 |
| rs2228145* | a | c | 0.360 | 0.0899 | 0.0042 | 1.2E-101 | 0.0027 | -0.0115 | 0.0009 | 0.0012 | 0.0635 | 0.0172 | 0.0018 | 1.114 | 0.0157 | 0.5719 |
| rs11264224 | a | c | 0.193 | 0.0465 | 0.0057 | 3.4E-16 | 0.0005 | -0.0078 | 0.0012 | 0.0004 | 0.0793 | 0.0243 | 0.0020 | 0.4576 | 0.0336 | 0.0652 |
| rs12059682 | t | c | 0.196 | -0.0441 | 0.0049 | 2.3E-19 | 0.0004 | -0.0055 | 0.0011 | 0.0002 | 0.0264 | 0.019 | 0.0002 | 0.3825 | 0.0299 | 0.0461 |
| rs34693607 | c | g | 0.184 | 0.0368 | 0.0057 | 1.1E-10 | 0.0003 | -0.0048 | 0.0012 | 0.0001 | 0.0541 | 0.0206 | 0.0009 | 0.1964 | 0.0293 | 0.0116 |

* Variants located within the *IL6R* gene.

† Beta coefficients correspond to 1-unit changes in the natural-log-transformed serum CRP (mg/L) levels,[1] 1-unit changes in the natural-log-transformed serum fibrinogen (Fg levels (g/L),[2] 1 SD units in circulating IL6 levels (pg/L),[3] and 1-unit changes in inverse-rank normalized soluble IL6 receptor (sIL6R) plasma levels[4] per copy increment in effect allele.

‡ R2 corresponds to the variance explained in each variable by the individuals SNPs and is calculated as $(2*MAF*(1-MAF)*b^2)/\sigma^2$.[5]

**Supplementary Table 2.** Biomarkers included in the analysis and sources/sample sizes.

| **Biomarker category** | **Source** | **N phenotypes** | **N individuals** | **Population (Ancestry)** | **Adjustments** |
|---|---|---|---|---|---|
| Hematological traits | UK Biobank & INTERVAL[6] | 36 | 173,480 | European | Age, sex |
| Cytokines/Growth factors | YES/FINRISK Studies[3] | 41 | 8,293 | Finnish | Age, sex, BMI |
| Glycemic traits | MAGIC[7] | 18 | up to 133,010 | European | Age, sex, BMI* |
| Hemodynamic traits | UK Biobank | 6 | 408,228 | White British | Age, sex |
| Infectious disease antigens | UK Biobank | 25 | 9,432 | White British | Age, sex |
| Serum lipids/metabolites | NMR GWAS[8] | 124 | 24,925 | European | Age, sex |
| Serum/urine biochemistry | UK Biobank | 25 | 353,579 | White British | Age, sex |
| Anthropometrics | GIANT | 91 | up to 322,154 | European | Age, sex |

* except for HbA1c: the analysis for HbA1c was performed in UK Biobank and we did not adjust for BMI

**Supplementary Table 3.** Characteristics of participants in the UK Biobank.

| Characteristics | Mean (SD)/ N (%) |
|---|---|
| N | 339,256 |
| Age, mean (SD) | 56.9 (8.0) |
| Female, N (%) | 182,110 (53.7) |
| BMI, kg/m2, mean (SD) | 27.4 (4.8) |
| SBP mmHg, mean (SD) | 136.3 (18.69) |
| On antihypertensive medications, N (%) | 74,507 (22.0) |
| LDL cholesterol, md/dl, mean (SD) | 138.1 (33.6) |
| On lipid-lowering medications, N (%) | 63,652 (18.8) |
| Hba1c, %, mean (SD) | 5.4 (2.7) |
| History of diabetes mellitus, N (%) | 17,331 (5.1) |
| Current smoker, N (%) | 34,024 (10.0) |

BMI: body mass index, HbA1c: glycated hemoglobin A1c, LDL: low-density lipoprotein, SBP: systolic blood pressure.

**Supplementary Table 4.** Characteristics of participants in the Penn Medicine Biobank (PMBB) and the BioMe Biobank.

| **Characteristics** | **PMBB** | **BioMe Biobank** |
|---|---|---|
| N | 10,244 | 9,054 |
| Age, mean (SD) | 71.0 (13.7) | 59.2 (19.2) |
| Female, N (%) | 3,515 (34.3) | 4,738 (52.3) |
| BMI, kg/m2, mean (SD) | 28.8 (6.4) | 27.1 (6.2) |
| SBP mmHg, mean (SD) | 126 (18) | 125 (22) |
| On antihypertensive medications, N (%) | 4,576 (44.7) | 4,339 (47.9) |
| LDL cholesterol, md/dl, mean (SD) | 91 (48) | 100 (37) |
| On lipid-lowering medications, N (%) | 6,842 (66.8) | 3,225 (35.6) |
| HbA1c, %, mean (SD) | 6.6 (2.8) | 6.3 (1.3) |
| History of diabetes mellitus, N (%) | 2,234 (21.8) | 915 (10.1) |
| Current smoker, N (%) | 517 (5.0) | 596 (6.6) |

BMI: body mass index, HbA1c: glycated hemoglobin A1c, LDL: low-density lipoprotein, SBP: systolic blood pressure.

**Supplementary Table 5.** Outcomes included in the analyses and outcomes excluded due to power.

| Diagnostic category | N Phenotypes | | | N cases | | |
|---|---|---|---|---|---|---|
| | **Total** | **Excluded due to power** | **Finally included** | **Median** | **Minimum** | **Maximum** |
| Infectious disease | 48 | 20 | 28 | 675 | 120 | 8439 |
| Neoplasms | 129 | 26 | 103 | 830 | 122 | 83,914 |
| Endocrine/ Metabolic | 102 | 45 | 57 | 458 | 123 | 35,924 |
| Hematopoietic | 40 | 17 | 23 | 368 | 147 | 10,086 |
| Mental disorders | 64 | 20 | 44 | 420 | 120 | 23,226 |
| Neurological | 68 | 22 | 46 | 433 | 120 | 32,094 |
| Sense organs | 104 | 34 | 70 | 534 | 120 | 31,844 |
| Circulatory system | 139 | 31 | 108 | 742 | 124 | 106,629 |
| Respiratory | 71 | 13 | 58 | 878 | 125 | 49,438 |
| Digestive | 143 | 25 | 118 | 1062 | 131 | 62,284 |
| Genitourinary | 140 | 28 | 112 | 1041 | 123 | 82,898 |
| Pregnancy complications | 28 | 9 | 19 | 773 | 124 | 7,518 |
| Dermatologic | 74 | 27 | 47 | 577 | 130 | 89,961 |
| Musculoskeletal | 109 | 28 | 81 | 826 | 120 | 59,848 |
| Congenital anomalies | 45 | 23 | 22 | 306 | 143 | 1454 |
| Symptoms | 27 | 6 | 21 | 1467 | 125 | 33,552 |
| Injuries/poisonings | 97 | 22 | 75 | 572 | 120 | 13,302 |

**Supplementary Table 6.** Detailed results for the effects of genetic downregulation of IL6 signaling on the examined clinical outcomes according to the PheCODE schema. Results are derived from inverse-variance weighted Mendelian randomization analyses.

(available as excel file)

**Supplementary Table 7.** Results of the sensitivity analyses for clinical outcomes.

| Diagnostic group | phecode | description | N cases | N controls | analysis | OR* | LCI | HCI | p-value | $p_{het}$ |
|---|---|---|---|---|---|---|---|---|---|---|
| neoplasms | 195 | Cancer, suspected or other | 879 | 248107 | IVW | 0.479 | 0.327 | 0.702 | 0.000164 | 0.9138 |
| | | | | | Weighted-median | 0.497 | 0.788 | 0.793 | 0.003 | |
| | | | | | IVW (3 SNPs) | 0.501 | 0.796 | 0.782 | 0.002 | |
| | 223 | Benign neoplasm of kidney and other urinary organs | 153 | 297014 | IVW | 4.238 | 1.720 | 10.43 | 0.001693 | 0.4499 |
| | | | | | Weighted-median | 5.700 | 0.562 | 17.62 | 0.003 | |
| | | | | | IVW (3 SNPs) | 4.848 | 0.586 | 13.83 | 0.003 | |
| endocrine/ metabolic | 250 | Diabetes mellitus | 16010 | 321754 | IVW | 0.813 | 0.739 | 0.895 | 2.11E-05 | 0.9124 |
| | | | | | Weighted-median | 0.842 | 0.943 | 0.945 | 0.003 | |
| | | | | | IVW (3 SNPs) | 0.837 | 0.945 | 0.936 | 0.002 | |
| | 250.2 | Type 2 diabetes | 15097 | 321754 | IVW | 0.804 | 0.730 | 0.886 | 9.55E-06 | 0.8882 |
| | | | | | Weighted-median | 0.822 | 0.943 | 0.922 | 0.001 | |
| | | | | | IVW (3 SNPs) | 0.815 | 0.945 | 0.911 | 0 | |
| neurological | 334.2 | Anterior horn cell disease | 165 | 304918 | IVW | 0.204 | 0.083 | 0.497 | 0.000466 | 0.7745 |
| | | | | | Weighted-median | 0.166 | 0.571 | 0.499 | 0.001 | |
| | | | | | IVW (3 SNPs) | 0.203 | 0.586 | 0.579 | 0.003 | |
| circulatory system | 396 | Abnormal heart sounds | 852 | 332812 | IVW | 0.476 | 0.323 | 0.702 | 0.00018 | 0.4216 |
| | | | | | Weighted-median | 0.424 | 0.776 | 0.697 | 0.001 | |
| | | | | | IVW (3 SNPs) | 0.531 | 0.794 | 0.833 | 0.006 | |
| | 411 | Ischemic Heart Disease | 24925 | 312241 | IVW | 0.831 | 0.767 | 0.898 | 3.47E-06 | 0.9304 |
| | | | | | Weighted-median | 0.823 | 0.952 | 0.907 | 0 | |
| | | | | | IVW (3 SNPs) | 0.829 | 0.954 | 0.909 | 0 | |
| | 411.3 | Angina pectoris | 13136 | 312241 | IVW | 0.756 | 0.681 | 0.840 | 1.6E-07 | 0.3083 |
| | | | | | Weighted-median | 0.742 | 0.935 | 0.847 | 0 | |
| | | | | | IVW (3 SNPs) | 0.785 | 0.940 | 0.886 | 0 | |
| | 411.4 | | 25105 | 312241 | IVW | 0.835 | 0.772 | 0.904 | 6.8E-06 | 0.9399 |

| | | | | | | | | | | |
|---|---|---|---|---|---|---|---|---|---|---|
| | | Coronary atherosclerosis | | | Weighted-median | 0.811 | 0.952 | 0.892 | 0 | |
| | | | | | IVW (3 SNPs) | 0.832 | 0.954 | 0.912 | 0 | |
| | 411.8 | Chronic ischemic heart disease, unspecified | 24872 | 312241 | IVW | 0.835 | 0.772 | 0.903 | 6.4E-06 | 0.943 |
| | | | | | Weighted-median | 0.827 | 0.952 | 0.910 | 0 | |
| | | | | | IVW (3 SNPs) | 0.833 | 0.954 | 0.913 | 0 | |
| | 420.21 | Acute pericarditis | 188 | 335549 | IVW | 3.815 | 1.699 | 8.565 | 0.001181 | 0.5433 |
| | | | | | Weighted-median | 4.016 | 0.607 | 10.70 | 0.005 | |
| | | | | | IVW (3 SNPs) | 3.021 | 0.620 | 7.722 | 0.021 | |
| | 442.11 | Abdominal aortic aneurysm | 726 | 330999 | IVW | 0.438 | 0.287 | 0.669 | 0.000136 | 0.7543 |
| | | | | | Weighted-median | 0.436 | 0.769 | 0.730 | 0.002 | |
| | | | | | IVW (3 SNPs) | 0.454 | 0.777 | 0.743 | 0.002 | |
| | 454.1 | Varicose veins, lower extremity | 9179 | 306018 | IVW | 0.802 | 0.712 | 0.903 | 0.000268 | 0.956 |
| | 454.1 | | | | Weighted-median | 0.796 | 0.930 | 0.917 | 0.002 | |
| | 454.1 | | | | IVW (3 SNPs) | 0.821 | 0.932 | 0.942 | 0.005 | |
| | 458.9 | Hypotension NOS | 2849 | 231296 | IVW | 1.512 | 1.221 | 1.871 | 0.000144 | 0.7129 |
| | | | | | Weighted-median | 1.332 | 0.876 | 1.728 | 0.031 | |
| | | | | | IVW (3 SNPs) | 1.437 | 0.882 | 1.838 | 0.004 | |
| respiratory | 495.2 | Asthma with exacerbation | 201 | 311025 | IVW | 3.621 | 1.652 | 7.935 | 0.001316 | 0.8971 |
| | | | | | Weighted-median | 4.032 | 0.618 | 10.37 | 0.004 | |
| | | | | | IVW (3 SNPs) | 3.356 | 0.630 | 8.306 | 0.009 | |
| | 506 | Empyema and pneumothorax | 927 | 328855 | IVW | 0.472 | 0.323 | 0.687 | 9.26E-05 | 0.945 |
| | | | | | Weighted-median | 0.489 | 0.792 | 0.772 | 0.002 | |
| | | | | | IVW (3 SNPs) | 0.512 | 0.800 | 0.793 | 0.003 | |
| | 519.2 | Respiratory complications | 242 | 288235 | IVW | 0.281 | 0.134 | 0.588 | 0.000748 | 0.463 |
| | | | | | Weighted-median | 0.386 | 0.634 | 0.943 | 0.037 | |
| | | | | | IVW (3 SNPs) | 0.415 | 0.646 | 0.976 | 0.044 | |
| digestive | 574.11 | Cholelithiasis with acute cholecystitis | 1227 | 323752 | IVW | 1.759 | 1.276 | 2.425 | 0.000558 | 0.7159 |
| | | | | | Weighted-median | 1.684 | 0.810 | 2.543 | 0.013 | |
| | | | | | IVW (3 SNPs) | 1.643 | 0.827 | 2.383 | 0.009 | |

| | | | | | | | | | | |
|---|---|---|---|---|---|---|---|---|---|---|
| | 574.12 | Cholelithiasis with other cholecystitis | 4489 | 323752 | IVW | 1.374 | 1.156 | 1.631 | 0.000295 | 0.9973 |
| | | | | | Weighted-median | 1.379 | 0.899 | 1.699 | 0.002 | |
| | | | | | IVW (3 SNPs) | 1.371 | 0.903 | 1.675 | 0.002 | |
| | 578.2 | Blood in stool | 2127 | 318771 | IVW | 1.524 | 1.195 | 1.945 | 0.000718 | 0.6635 |
| | | | | | Weighted-median | 1.345 | 0.855 | 1.829 | 0.059 | |
| | | | | | IVW (3 SNPs) | 1.506 | 0.866 | 1.998 | 0.004 | |
| genitourinary | 591 | Urinary tract infection | 10044 | 313404 | IVW | 1.254 | 1.117 | 1.409 | 0.000139 | 0.7539 |
| | | | | | Weighted-median | 1.232 | 0.930 | 1.421 | 0.004 | |
| | | | | | IVW (3 SNPs) | 1.252 | 0.933 | 1.433 | 0.001 | |
| | 597 | Other disorders of urethra and urinary tract | 709 | 326499 | IVW | 2.029 | 1.331 | 3.095 | 0.001017 | 0.6111 |
| | | | | | Weighted-median | 2.007 | 0.766 | 3.384 | 0.009 | |
| | | | | | IVW (3 SNPs) | 1.947 | 0.779 | 3.177 | 0.008 | |
| | 626.8 | Infertility, female | 1146 | 312766 | IVW | 1.787 | 1.274 | 2.507 | 0.000782 | 0.9594 |
| | | | | | Weighted-median | 1.762 | 0.811 | 2.658 | 0.007 | |
| | | | | | IVW (3 SNPs) | 1.817 | 0.819 | 2.686 | 0.003 | |
| | 627.3 | Postmenopausal atrophic vaginitis | 908 | 312766 | IVW | 2.119 | 1.460 | 3.073 | 7.6E-05 | 0.8096 |
| | | | | | Weighted-median | 2.060 | 0.792 | 3.251 | 0.002 | |
| | | | | | IVW (3 SNPs) | 2.140 | 0.803 | 3.291 | 0.001 | |
| dermatologic | 681.3 | Cellulitis and abscess of arm/hand | 4502 | 328919 | IVW | 1.352 | 1.138 | 1.605 | 0.000585 | 0.1798 |
| | | | | | Weighted-median | 1.457 | 0.896 | 1.806 | 0.001 | |
| | | | | | IVW (3 SNPs) | 1.379 | 0.903 | 1.685 | 0.002 | |
| | 681.5 | Cellulitis and abscess of leg, except foot | 4504 | 328919 | IVW | 1.336 | 1.125 | 1.587 | 0.000953 | 0.2342 |
| | | | | | Weighted-median | 1.430 | 0.897 | 1.771 | 0.001 | |
| | | | | | IVW (3 SNPs) | 1.374 | 0.903 | 1.679 | 0.002 | |
| | 681.6 | Cellulitis and abscess of foot, toe | 4469 | 328919 | IVW | 1.342 | 1.129 | 1.594 | 0.000864 | 0.2097 |
| | | | | | Weighted-median | 1.488 | 0.897 | 1.843 | 0 | |
| | | | | | IVW (3 SNPs) | 1.380 | 0.903 | 1.686 | 0.002 | |
| | 689 | | 33216 | 304955 | IVW | 1.133 | 1.062 | 1.208 | 0.000147 | 0.4973 |
| | | | | | Weighted-median | 1.119 | 0.961 | 1.210 | 0.005 | |

| | | | | | | | | | | |
|---|---|---|---|---|---|---|---|---|---|---|
| | | Disorder of skin and subcutaneous tissue NOS | | | IVW (3 SNPs) | 1.130 | 0.963 | 1.218 | 0.001 | |
| | 695.9 | Unspecified erythematous condition | 380 | 333072 | IVW | 2.903 | 1.629 | 5.171 | 0.000294 | 0.5892 |
| | | | | | Weighted-median | 2.447 | 0.697 | 4.958 | 0.013 | |
| | | | | | IVW (3 SNPs) | 2.160 | 0.711 | 4.214 | 0.024 | |
| | 703.1 | Ingrowing nail | 785 | 332779 | IVW | 2.543 | 1.692 | 3.821 | 7E-06 | 0.0545 |
| | | | | | Weighted-median | 1.812 | 0.769 | 3.029 | 0.023 | |
| | | | | | IVW (3 SNPs) | 2.093 | 0.787 | 3.349 | 0.002 | |
| | 707.1 | Decubitus ulcer | 211 | 337065 | IVW | 0.279 | 0.128 | 0.612 | 0.001437 | 0.7253 |
| | | | | | Weighted-median | 0.290 | 0.616 | 0.749 | 0.011 | |
| | | | | | IVW (3 SNPs) | 0.298 | 0.626 | 0.745 | 0.01 | |
| | 939 | Atopic/contact dermatitis due to other or unspecified | 1696 | 333163 | IVW | 2.137 | 1.619 | 2.822 | 8.96E-08 | 0.3634 |
| | | | | | Weighted-median | 2.351 | 0.837 | 3.337 | 0 | |
| | | | | | IVW (3 SNPs) | 2.081 | 0.849 | 2.872 | 0 | |
| musculoskeletal | 733.2 | Cyst of bone | 250 | 323543 | IVW | 3.803 | 1.856 | 7.798 | 0.000264 | 0.2888 |
| | | | | | Weighted-median | 2.796 | 0.638 | 6.748 | 0.022 | |
| | | | | | IVW (3 SNPs) | 2.626 | 0.655 | 6.022 | 0.023 | |
| | 740.2 | Osteoarthrosis, generalized | 480 | 314842 | IVW | 2.325 | 1.391 | 3.886 | 0.001286 | 0.3217 |
| | | | | | Weighted-median | 2.008 | 0.719 | 3.829 | 0.034 | |
| | | | | | IVW (3 SNPs) | 2.490 | 0.739 | 4.502 | 0.003 | |
| | 742.1 | Loose body in joint | 276 | 307651 | IVW | 0.328 | 0.163 | 0.657 | 0.001672 | 0.4904 |
| | | | | | Weighted-median | 0.363 | 0.655 | 0.832 | 0.017 | |
| | | | | | IVW (3 SNPs) | 0.396 | 0.664 | 0.883 | 0.024 | |

The odds ratios correspond to the CRP-decreasing effects of tocilizumab (genetically downregulated IL-6 signaling.

IVW: inverse-variance weighted.

**Supplementary Table 8.** Results of the meta-analysis of the Mendelian randomization effects derived from the UK Biobank, Penn Medicine Biobank, and the BioMe Biobank.

| phecode | description | biobank | N cases | N controls | OR | LCI | HCI | P-value | P-heterogeneity* |
|---|---|---|---|---|---|---|---|---|---|
| 195 | Cancer, suspected or other | PMBB | 247 | 8887 | 0.928 | 0.441 | 1.954 | 0.84400 | 0.9131 |
| | | BioMe | 45 | 9009 | 1.252 | 0.225 | 6.966 | 0.79800 | 0.9371 |
| | | UKB | 879 | 248107 | 0.479 | 0.327 | 0.702 | 0.00016 | 0.9138 |
| | | Meta-analysis | 924 | 257116 | 0.648 | 0.375 | 1.12 | 0.12000 | 0.197 |
| 223 | Benign neoplasm of kidney and other urinary organs | UKB | 153 | 297014 | 4.238 | 1.72 | 10.43 | 0.00169 | 0.4499 |
| | | Meta-analysis | | | not possible | | | | |
| **250** | **Diabetes mellitus** | PMBB | 2534 | 6414 | 0.855 | 0.651 | 1.122 | 0.25800 | 0.7056 |
| | | BioMe | 209 | 8845 | 0.908 | 0.406 | 2.035 | 0.81500 | 0.4842 |
| | | UKB | 16010 | 321754 | 0.813 | 0.739 | 0.895 | 2.1E-05 | 0.9124 |
| | | **Meta-analysis** | **18753** | **337013** | **0.819** | **0.748** | **0.895** | **1.2E-05** | **0.914** |
| **250.2** | **Type 2 diabetes** | PMBB | 2119 | 6414 | 0.793 | 0.593 | 1.061 | 0.11800 | 0.5516 |
| | | BioMe | 727 | 8327 | 0.798 | 0.506 | 1.256 | 0.32900 | 0.2641 |
| | | UKB | 15097 | 321754 | 0.804 | 0.73 | 0.886 | 9.6E-06 | 0.8882 |
| | | **Meta-analysis** | **17943** | **336495** | **0.803** | **0.734** | **0.878** | **1.8E-06** | **0.996** |
| 334.2 | Anterior horn cell disease | UKB | 165 | 304918 | 0.204 | 0.083 | 0.497 | 0.00047 | 0.7745 |
| | | Meta-analysis | | | not possible | | | | |
| **396** | **Abnormal heart sounds** | PMBB | 131 | 5729 | 0.435 | 0.155 | 1.224 | 0.11500 | 0.8857 |
| | | BioMe | 105 | 8949 | 0.319 | 0.103 | 0.989 | 0.04800 | 0.8557 |
| | | UKB | 852 | 332812 | 0.476 | 0.323 | 0.702 | 0.00018 | 0.4216 |
| | | **Meta-analysis** | **1088** | **347490** | **0.454** | **0.321** | **0.642** | **7.8E-06** | **0.804** |
| 411 | Ischemic Heart Disease | PMBB | 4370 | 4702 | 0.952 | 0.73 | 1.242 | 0.71700 | 0.7674 |
| | | BioMe | 17 | 9037 | 33.699 | 1.947 | 583.631 | 0.01600 | 0.4656 |
| | | UKB | 24925 | 312241 | 0.831 | 0.767 | 0.898 | 3.5E-06 | 0.9304 |
| | | Meta-analysis | 29312 | 325980 | 0.928 | 0.659 | 1.306 | 0.66800 | 0.025 |

| | | | | | | | | | | |
|---|---|---|---|---|---|---|---|---|---|---|
| **411.3** | **Angina pectoris** | PMBB | 479 | 4702 | 0.976 | 0.555 | 1.718 | 0.93300 | | 0.6187 |
| | | BioMe | 99 | 8955 | 0.989 | 0.304 | 3.218 | 0.98600 | | 0.8247 |
| | | UKB | 13136 | 312241 | 0.756 | 0.681 | 0.84 | 1.6E-07 | | 0.3083 |
| | | **Meta-analysis** | **13714** | **325898** | **0.764** | **0.689** | **0.847** | **2.9E-07** | | **0.623** |
| **411.4** | **Coronary atherosclerosis** | PMBB | 3997 | 4702 | 0.909 | 0.691 | 1.195 | 0.49600 | | 0.5362 |
| | | BioMe | 732 | 8322 | 1.038 | 0.643 | 1.676 | 0.87900 | | 0.8323 |
| | | UKB | 25105 | 312241 | 0.835 | 0.772 | 0.904 | 6.8E-06 | | 0.9399 |
| | | **Meta-analysis** | **29834** | **325265** | **0.845** | **0.784** | **0.911** | **1.0E-05** | | **0.587** |
| **411.8** | **Chronic ischemic heart disease, unspecified** | PMBB | 899 | 4702 | 0.968 | 0.616 | 1.52 | 0.88700 | | 0.6597 |
| | | BioMe | 35 | 9019 | 0.664 | 0.08 | 5.485 | 0.70400 | | 0.0436 |
| | | UKB | 24872 | 312241 | 0.835 | 0.772 | 0.903 | 6.4E-06 | | 0.943 |
| | | **Meta-analysis** | **25806** | **325962** | **0.838** | **0.776** | **0.906** | **7.5E-06** | | **0.801** |
| 420.21 | Acute pericarditis | UKB | 188 | 335549 | 3.815 | 1.699 | 8.565 | 0.00118 | | 0.5433 |
| | | Meta-analysis | | | not possible | | | | | |
| **442.11** | **Abdominal aortic aneurysm** | PMBB | 388 | 6602 | 0.63 | 0.339 | 1.17 | 0.14300 | | 0.0652 |
| | | BioMe | 227 | 8827 | 0.757 | 0.33 | 1.737 | 0.51100 | | 0.115 |
| | | UKB | 726 | 330999 | 0.438 | 0.287 | 0.669 | 0.00014 | | 0.7543 |
| | | **Meta-analysis** | **1341** | **346428** | **0.525** | **0.38** | **0.724** | **8.5E-05** | | **0.411** |
| **454.1** | **Varicose veins, lower extremity** | PMBB | 134 | 6559 | 0.864 | 0.315 | 2.375 | 0.77800 | | 0.908 |
| | | BioMe | 53 | 9001 | 0.547 | 0.114 | 2.624 | 0.45100 | | 0.6765 |
| | | UKB | 9179 | 306018 | 0.802 | 0.712 | 0.903 | 0.00027 | | 0.956 |
| | | **Meta-analysis** | **9366** | **321578** | **0.801** | **0.712** | **0.901** | **0.00022** | | **0.883** |
| **458.9** | **Hypotension NOS** | PMBB | 191 | 7749 | 1.379 | 0.58 | 3.28 | 0.46600 | | 0.2336 |
| | | BioMe | 33 | 9021 | 4.593 | 0.545 | 38.703 | 0.16100 | | 0.6607 |
| | | UKB | 2849 | 231296 | 1.512 | 1.221 | 1.871 | 0.00014 | | 0.7129 |
| | | **Meta-analysis** | **3073** | **248066** | **1.52** | **1.237** | **1.868** | **6.9E-05** | | **0.581** |
| 495.2 | Asthma with exacerbation | PMBB | 64 | 7586 | 2.452 | 0.562 | 10.706 | 0.23300 | | 0.3129 |

| | | | | | | | | | |
|---|---|---|---|---|---|---|---|---|---|
| | | BioMe | 100 | 8954 | 0.259 | 0.082 | 0.819 | 0.02100 | 0.9984 |
| | | UKB | 201 | 311025 | 3.621 | 1.652 | 7.935 | 0.00132 | 0.8971 |
| | | Meta-analysis | 365 | 327565 | 1.335 | 0.243 | 7.335 | 0.74000 | 0.001 |
| 506 | Empyema and pneumothorax | PMBB | 79 | 7614 | 2.322 | 0.647 | 8.336 | 0.19600 | 0.7446 |
| | | BioMe | 9 | 9045 | 0.086 | 0.002 | 3.696 | 0.20100 | 0.9345 |
| | | UKB | 927 | 328855 | 0.472 | 0.323 | 0.687 | 9.3E-05 | 0.945 |
| | | Meta-analysis | 1015 | 345514 | 0.709 | 0.178 | 2.815 | 0.62500 | 0.041 |
| 519.2 | Respiratory complications | UKB | 242 | 288235 | 0.281 | 0.134 | 0.588 | 0.00075 | 0.463 |
| | | Meta-analysis | | | not possible | | | | |
| 574.11 | Cholelithiasis with acute cholecystitis | UKB | 1227 | 323752 | 1.759 | 1.276 | 2.425 | 0.00056 | 0.7159 |
| | | Meta-analysis | | | not possible | | | | |
| | | UKB | 4489 | 323752 | 1.374 | 1.156 | 1.631 | 0.00029 | 0.9973 |
| | | Meta-analysis | | | not possible | | | | |
| **591** | **Urinary tract infection** | PMBB | 541 | 8192 | 1.094 | 0.653 | 1.834 | 0.73200 | 0.5409 |
| | | BioMe | 333 | 8721 | 0.901 | 0.473 | 1.714 | 0.75000 | 0.8044 |
| | | UKB | 10044 | 313404 | 1.254 | 1.117 | 1.409 | 0.00014 | 0.7539 |
| | | **Meta-analysis** | **10918** | **330317** | **1.234** | **1.103** | **1.379** | **0.00022** | **0.549** |
| **597** | **Other disorders of urethra and urinary tract** | PMBB | 67 | 9452 | 1.162 | 0.283 | 4.771 | 0.83500 | 0.4613 |
| | | UKB | 709 | 326499 | 2.029 | 1.331 | 3.095 | 0.00102 | 0.6111 |
| | | **Meta-analysis** | **776** | **335951** | **1.938** | **1.294** | **2.904** | **0.00100** | **0.458** |
| **626.8** | **Infertility, female** | PMBB | 34 | 9511 | 1.538 | 0.145 | 16.311 | 0.72100 | 0.2804 |
| | | BioMe | 18 | 9036 | 1.286 | 0.084 | 19.618 | 0.85600 | 0.968 |
| | | UKB | 1146 | 312766 | 1.787 | 1.274 | 2.507 | 0.00078 | 0.9594 |
| | | **Meta-analysis** | **1198** | **331313** | **1.773** | **1.271** | **2.472** | **0.00100** | **0.966** |
| 627.3 | Postmenopausal atrophic vaginitis | PMBB | 105 | 9511 | 2.895 | 0.913 | 9.184 | 0.07100 | 0.3153 |
| | | BioMe | 115 | 8939 | 0.689 | 0.232 | 2.048 | 0.50300 | 0.398 |

| | | | | | | | | | |
|---|---|---|---|---|---|---|---|---|---|
| | | UKB | 908 | 312766 | 2.119 | 1.46 | 3.073 | 7.6E-05 | 0.8096 |
| | | Meta-analysis | 1128 | 331216 | 1.727 | 0.862 | 3.459 | 0.12300 | 0.126 |
| **681.3** | **Cellulitis and abscess of arm/hand** | PMBB | 25 | 9033 | 0.565 | 0.057 | 5.651 | 0.62700 | 0.7388 |
| | | BioMe | 7 | 9047 | 1.638 | 0.024 | 110.233 | 0.81800 | 0.6541 |
| | | UKB | 4502 | 328919 | 1.352 | 1.138 | 1.605 | 0.00058 | 0.1798 |
| | | **Meta-analysis** | **4534** | **346999** | **1.346** | **1.134** | **1.597** | **0.00100** | **0.757** |
| 681.5 | Cellulitis and abscess of leg, except foot | PMBB | 139 | 9033 | 2.561 | 0.954 | 6.874 | 0.06200 | 0.3909 |
| | | BioMe | 74 | 8980 | 0.531 | 0.137 | 2.063 | 0.36000 | 0.3196 |
| | | UKB | 4504 | 328919 | 1.336 | 1.125 | 1.587 | 0.00095 | 0.2342 |
| | | Meta-analysis | 4717 | 346932 | 1.358 | 0.769 | 2.397 | 0.29100 | 0.179 |
| 681.6 | Cellulitis and abscess of foot, toe | BioMe | 6 | 9048 | 0.002 | 0 | 0.414 | 0.02200 | 0.5666 |
| | | UKB | 4469 | 328919 | 1.342 | 1.129 | 1.594 | 0.00086 | 0.2097 |
| | | Meta-analysis | 4475 | 337967 | 0.094 | 0 | 46.49 | 0.45500 | 0.016 |
| 689 | Disorder of skin and subcutaneous tissue NOS | PMBB | 116 | 9722 | 1.002 | 0.34 | 2.958 | 0.99700 | 0.7415 |
| | | BioMe | 34 | 9020 | 29.949 | 3.94 | 227.514 | 0.00100 | 0.4327 |
| | | UKB | 33216 | 304955 | 1.133 | 1.062 | 1.208 | 0.00015 | 0.4973 |
| | | Meta-analysis | 33366 | 323697 | 2.175 | 0.601 | 7.868 | 0.23600 | 0.007 |
| **695.9** | **Unspecified erythematous condition** | PMBB | 23 | 8787 | 2.94 | 0.269 | 32.146 | 0.37700 | 0.4341 |
| | | UKB | 380 | 333072 | 2.903 | 1.629 | 5.171 | 0.00029 | 0.5892 |
| | | **Meta-analysis** | **403** | **341859** | **2.905** | **1.657** | **5.093** | **0.00020** | **0.992** |
| **707.1** | **Decubitus ulcer** | PMBB | 173 | 9683 | 0.684 | 0.279 | 1.68 | 0.40800 | 0.8049 |
| | | BioMe | 31 | 9023 | 0.36 | 0.042 | 3.087 | 0.35100 | 0.4083 |
| | | UKB | 211 | 337065 | 0.279 | 0.128 | 0.612 | 0.00144 | 0.7253 |
| | | **Meta-analysis** | **415** | **355771** | **0.41** | **0.242** | **0.715** | **0.00128** | **0.334** |
| 733.2 | Cyst of bone | UKB | 1696 | 333163 | 3.803 | 1.856 | 7.798 | 0.00026 | 0.2888 |

| | | | | | | | | | |
|---|---|---|---|---|---|---|---|---|---|
| | | Meta-analysis | | | not possible | | | | |
| 740.2 | Osteoarthrosis, generalized | PMBB | 193 | 8268 | 2.093 | 0.9 | 4.862 | 0.08600 | 0.5063 |
| | | BioMe | 72 | 8982 | 0.342 | 0.086 | 1.364 | 0.12800 | 0.5894 |
| | | UKB | 250 | 323543 | 2.325 | 1.391 | 3.886 | 0.00129 | 0.3217 |
| | | Meta-analysis | 515 | 340793 | 1.449 | 0.591 | 3.552 | 0.41700 | 0.038 |
| 742.1 | Loose body in joint | UKB | 480 | 314842 | 0.328 | 0.163 | 0.657 | 0.00167 | 0.4904 |
| | | Meta-analysis | | | not possible | | | | |
| **939** | **Atopic/contact dermatitis due to other or unspecified** | PMBB | 411 | 8518 | 1.781 | 0.995 | 3.19 | 0.05200 | 0.4581 |
| | | BioMe | 338 | 8716 | 1.511 | 1.282 | 2.927 | 0.22100 | 0.9578 |
| | | UKB | 276 | 307651 | 2.137 | 1.619 | 2.822 | 9.0E-08 | 0.3634 |
| | | **Meta-analysis** | **1025** | **324885** | **1.899** | **1.533** | **2.353** | **4.4E-09** | **0.383** |

In individual study estimates for Penn Medicine Biobank (PMBB), BioMe Biobank, and UK Biobank (UKB) are derived from inverse-variance weighted Mendelian randomization analyses. The meta-analysis effects are derived from random-effect meta-models (DerSimonian-Laird).

In **bold** are the results from the meta-analyses that were still statistically significant at the original FDR<0.05 threshold.

* P-values for heterogeneity refer to between-SNP heterogeneity in the inverse-variance weighted Mendelian randomization analyses for the individual biobanks (PMBB, BioMe, UKB) and to between-biobank heterogeneity in the random-effect meta-analysis for the overall effect. Both are derived from the Cochran Q statistic.

**Supplementary Table 9.** Detailed results for the effects of genetic downregulation of IL6 signaling on the examined biomarkers. Results are derived from inverse-variance weighted Mendelian randomization analyses.

(available as excel file)

**Supplementary Table 10.** Results of the sensitivity analyses for biomarkers.

| Biomarker category | Biomarker | analysis | BETA* | LCI | HCI | pval | p-heter |
|---|---|---|---|---|---|---|---|
| Hematological traits | Mean corpuscular hemoglobin | IVW | 0.119 | 0.09 | 0.148 | 1.64E-15 | 0.6482 |
| | | weighted median | 0.108 | 0.073 | 0.144 | 4.6E-09 | |
| | | IVW (3 SNPs) | 0.123 | 0.089 | 0.157 | 1.68E-12 | |
| | Monocyte percentage of white cells | IVW | 0.112 | 0.083 | 0.141 | 5.6E-14 | 0.3383 |
| | | weighted median | 0.122 | 0.086 | 0.158 | 3.82E-11 | |
| | | IVW (3 SNPs) | 0.122 | 0.089 | 0.157 | 2.08E-12 | |
| | Red cell distribution width | IVW | -0.112 | -0.14 | -0.082 | 5.6E-14 | 0.9437 |
| | | weighted median | -0.106 | -0.142 | -0.071 | 3.34E-09 | |
| | | IVW (3 SNPs) | -0.105 | -0.138 | -0.071 | 5.23E-10 | |
| | Mean corpuscular volume | IVW | 0.103 | 0.075 | 0.133 | 3.26E-12 | 0.5922 |
| | | weighted median | 0.093 | 0.058 | 0.128 | 2.33E-07 | |
| | | IVW (3 SNPs) | 0.103 | 0.07 | 0.137 | 1.12E-09 | |
| | Hemoglobin concentration | IVW | 0.103 | 0.074 | 0.132 | 4.18E-12 | 0.9507 |
| | | weighted median | 0.103 | 0.069 | 0.139 | 7.88E-09 | |
| | | IVW (3 SNPs) | 0.102 | 0.069 | 0.135 | 1.97E-09 | |
| | Granulocyte percentage of myeloid white cells | IVW | -0.1 | -0.128 | -0.07 | 2.23E-11 | 0.4364 |
| | | weighted median | -0.105 | -0.142 | -0.069 | 1.05E-08 | |
| | | IVW (3 SNPs) | -0.111 | -0.145 | -0.078 | 1.75E-10 | |
| | Hematocrit | IVW | 0.083 | 0.054 | 0.112 | 7.21E-09 | 0.8764 |
| | | weighted median | 0.083 | 0.049 | 0.118 | 1.89E-06 | |
| | | IVW (3 SNPs) | 0.076 | 0.042 | 0.109 | 7.29E-06 | |
| | Monocyte count | IVW | 0.084 | 0.055 | 0.113 | 1.56E-08 | 0.6517 |
| | | weighted median | 0.089 | 0.054 | 0.124 | 7.69E-07 | |
| | | IVW (3 SNPs) | 0.089 | 0.055 | 0.122 | 1.59E-07 | |
| | Mean corpuscular hemoglobin concentration | IVW | 0.066 | 0.037 | 0.093 | 4.85E-06 | 0.5289 |

| | | | | | | | |
|---|---|---|---|---|---|---|---|
| | | weighted median | 0.071 | 0.038 | 0.105 | 3.82E-05 | |
| | | IVW (3 SNPs) | 0.083 | 0.05 | 0.114 | 5.73E-07 | |
| | Platelet distribution width | IVW | -0.056 | -0.084 | -0.026 | 0.000171 | 0.0358 |
| | | weighted median | -0.059 | -0.094 | -0.023 | 0.001126 | |
| | | IVW (3 SNPs) | -0.066 | -0.099 | -0.033 | 0.000105 | |
| Cytokines/Growth factors | Interleukin-6 | IVW | 0.308 | 0.177 | 0.439 | 3.78E-06 | 0.1246 |
| | | weighted median | 0.328 | 0.16 | 0.498 | 0.000139 | |
| | | IVW (3 SNPs) | 0.255 | 0.103 | 0.407 | 0.000974 | |
| | CTACK (Cutaneous T-Cell Attracting Chemokine) | IVW | 0.406 | 0.213 | 0.599 | 3.71E-05 | 0.1648 |
| | | weighted median | 0.466 | 0.214 | 0.718 | 0.000292 | |
| | | IVW (3 SNPs) | 0.369 | 0.144 | 0.594 | 0.001309 | |
| | Interleukin-4 | IVW | 0.259 | 0.128 | 0.389 | 0.000106 | 0.1108 |
| | | weighted median | 0.245 | 0.077 | 0.415 | 0.004358 | |
| | | IVW (3 SNPs) | 0.161 | 0.01 | 0.313 | 0.037617 | |
| Glycemic traits | Glycated haemoglobin (HbA1c) | IVW | -0.067 | -0.084 | -0.05 | 1.3E-14 | 0.3882 |
| | | weighted median | -0.079 | -0.102 | -0.057 | 1.59E-11 | |
| | | IVW (3 SNPs) | -0.066 | -0.087 | -0.046 | 1.12E-10 | |
| Infectious disease antigens | HPV-16 type 2 | IVW | -1.306 | -1.931 | -0.677 | 4.66E-05 | 0.0877 |
| | | weighted median | -1.483 | -2.263 | -0.699 | 0.000208 | |
| | | IVW (3 SNPs) | -1.604 | -2.343 | -0.872 | 1.83E-05 | |
| Serum lipids/ metabolites | Apolipoprotein A | IVW | 0.07 | 0.052 | 0.087 | 6.38E-14 | 0.0009 |
| | | weighted median | 0.062 | 0.037 | 0.087 | 4.61E-07 | |
| | | IVW (3 SNPs) | 0.052 | 0.031 | 0.071 | 4.42E-07 | |
| | HDL cholesterol | IVW | 0.039 | 0.022 | 0.056 | 5.93E-06 | 0.1182 |
| | | weighted median | 0.034 | 0.011 | 0.058 | 0.00358 | |
| | | IVW (3 SNPs) | 0.028 | 0.008 | 0.047 | 0.008049 | |
| | Total cholesterol | IVW | 0.039 | 0.022 | 0.057 | 1.89E-05 | 0.466 |
| | | weighted median | 0.048 | 0.024 | 0.071 | 8.97E-05 | |
| | | IVW (3 SNPs) | 0.04 | 0.02 | 0.061 | 0.000204 | |

| | | | | | | | |
|---|---|---|---|---|---|---|---|
| Serum/urine biochemistry | C-reactive protein | IVW | -0.506 | -0.543 | -0.468 | 1.8E-156 | 0.7065 |
| | | weighted median | -0.528 | -0.574 | -0.481 | 4.8E-106 | |
| | | IVW (3 SNPs) | -0.521 | -0.564 | -0.476 | 9.7E-118 | |
| | Alkaline phosphatase | IVW | -0.161 | -0.182 | -0.142 | 7.34E-51 | 0.0014 |
| | | weighted median | -0.185 | -0.211 | -0.16 | 2.9E-47 | |
| | | IVW (3 SNPs) | -0.188 | -0.212 | -0.163 | 2.81E-48 | |
| | Albumin | IVW | 0.107 | 0.087 | 0.127 | 2.48E-25 | 0.0215 |
| | | weighted median | 0.11 | 0.083 | 0.137 | 2.26E-15 | |
| | | IVW (3 SNPs) | 0.101 | 0.079 | 0.124 | 3.4E-19 | |
| | SHBG | IVW | 0.091 | 0.072 | 0.109 | 8.12E-23 | 0.0003 |
| | | weighted median | 0.082 | 0.057 | 0.106 | 2.02E-10 | |
| | | IVW (3 SNPs) | 0.075 | 0.054 | 0.096 | 1.98E-13 | |
| | Total bilirubin | IVW | 0.06 | 0.043 | 0.079 | 5.53E-11 | 0.00001 |
| | | weighted median | 0.072 | 0.048 | 0.097 | 1.7E-08 | |
| | | IVW (3 SNPs) | 0.054 | 0.034 | 0.075 | 8.8E-08 | |
| | Calcium | IVW | 0.046 | 0.027 | 0.065 | 2.81E-06 | 0.0291 |
| | | weighted median | 0.024 | 0.002 | 0.046 | 0.036528 | |
| | | IVW (3 SNPs) | 0.046 | 0.024 | 0.068 | 5.23E-05 | |
| | Cystatin C | IVW | -0.035 | -0.051 | -0.019 | 2.14E-05 | 0.1487 |
| | | weighted median | -0.026 | -0.046 | -0.007 | 0.010772 | |
| | | IVW (3 SNPs) | -0.03 | -0.05 | -0.012 | 0.001589 | |

IVW: inverse-variance weighted.

**Supplementary Figure 1.** Overview of the genomic region that was screened for identification of genetic instruments for IL6 signaling and precise location of the identified single nucleotide polymorphisms (SNPs).

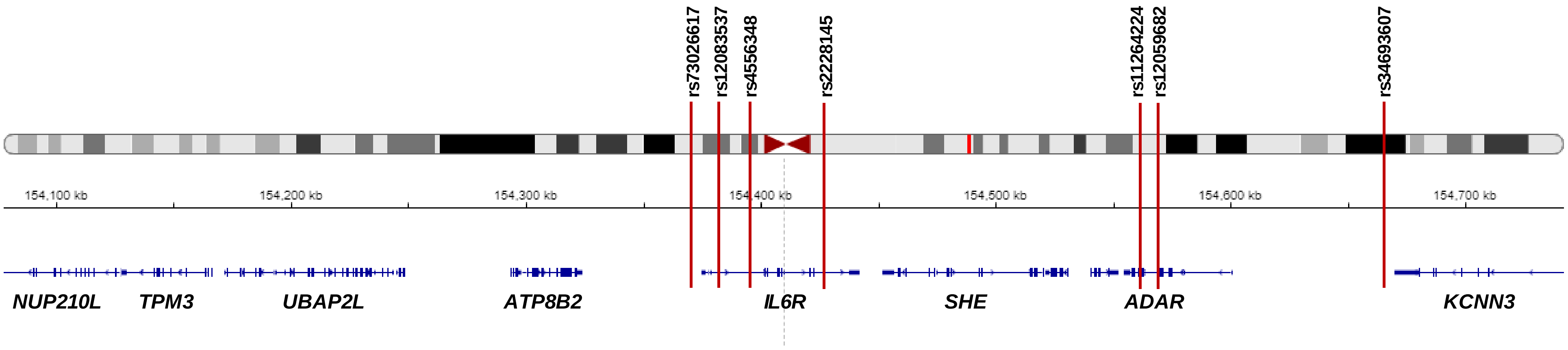


The region was selected as 300 kB upstream or downstream from the *IL6R* gene according to GRCh37/hg19 (chr1: 154,077,669-154,741,926). Gene name annotations are presented according to GENCODE (version 28) and accessed through the Integrative Genomics Viewer (https://igv.org/app/).

**Supplementary Figure 2.** Associations of the genetic variants used as instruments for IL6 signaling downregulation with upstream and downstream molecules in the IL6 signaling cascade.

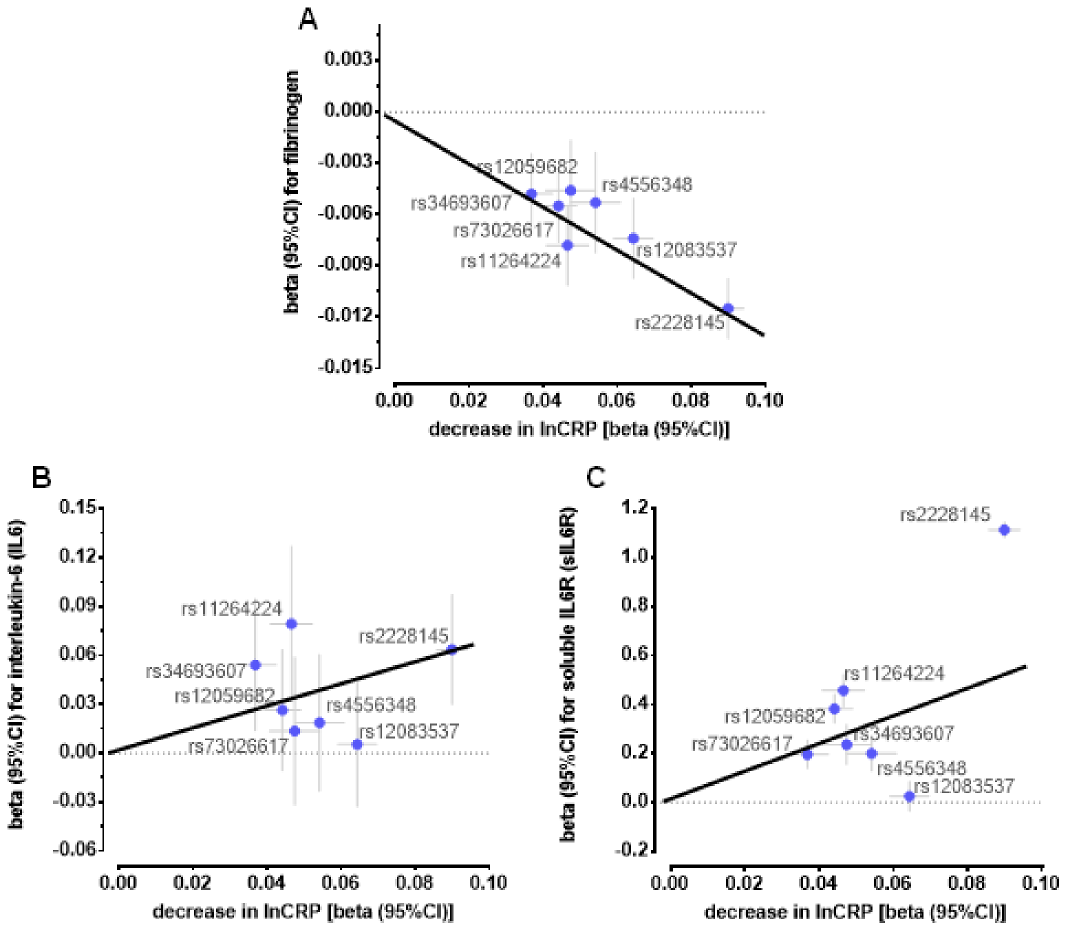


The black lines correspond to the Mendelian randomization estimates as derived from inverse-variance-weighted fixed-effects analyses (A: beta: -0.124, 95%CI: -0.141 to -0.108, $p=3.5x10^{-54}$; B: beta: 0.601, 95%CI: 0.346 to 0.856, $p=3.8x10^{-6}$; C: beta: 5.318, 95%CI: 4.736 to 5.900, $p=1.1x10^{-71}$).

Beta coefficients correspond to decreases in natural log-transformed serum C-reactive protein (CRP) levels, changes in natural log-transformed serum fibrinogen levels, SD changes in circulating IL6 levels, and changes in inverse-rank transformed sIL6R plasma levels.

**Supplementary Figure 3.** Power calculations for the clinical endpoints.

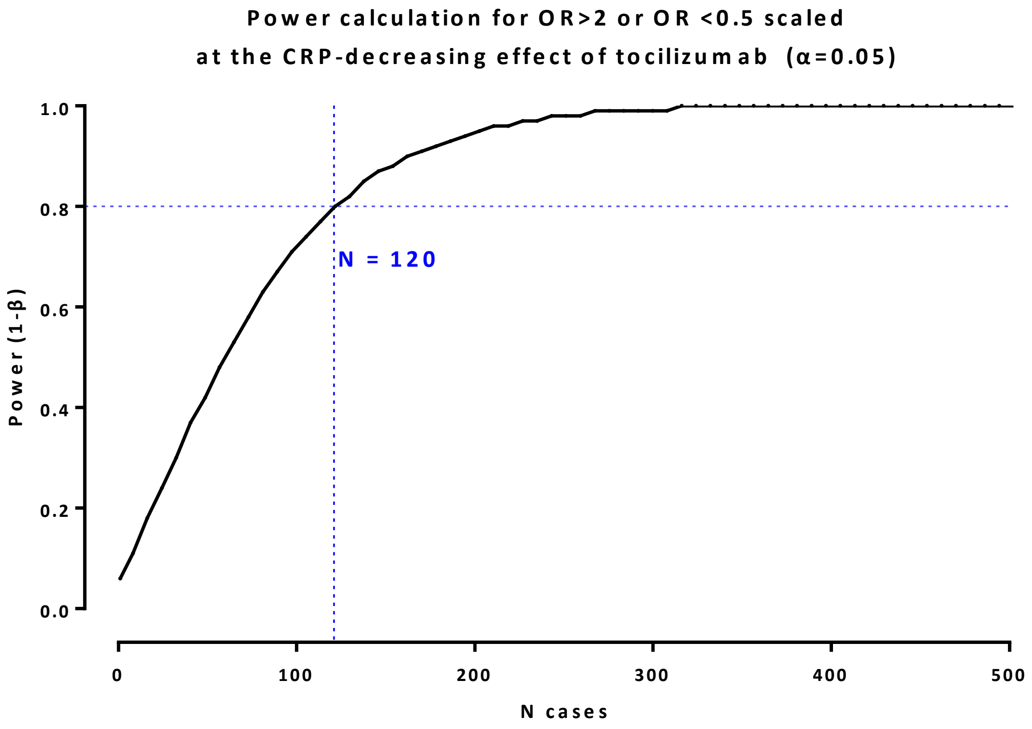

**Supplementary Figure 4.** Flowchart of the significant associations for biomarkers.

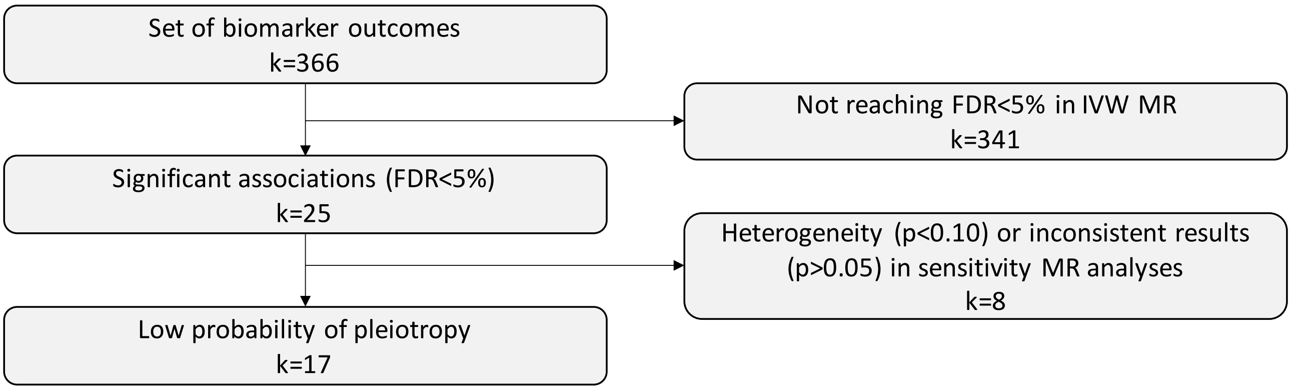


*weighted median, IVW restricted to 3 SNPs within *IL6R*

**Supplementary Figure 5.** Associations between genetically downregulated IL6 signaling and main lipid biomarkers.

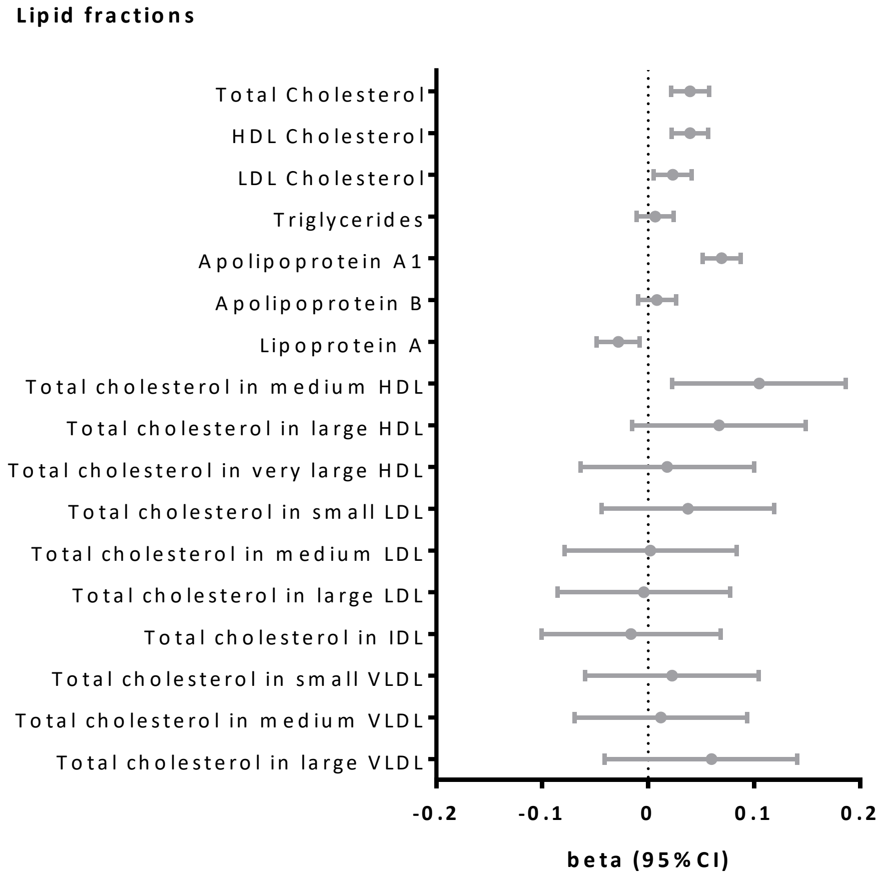


The beta coefficients correspond to the CRP-decreasing effect of tocilizumab.

**Supplementary References**


1. Ligthart S, Vaez A, Vosa U, Stathopoulou MG, de Vries PS, Prins BP, Van der Most PJ, Tanaka T, Naderi E, Rose LM, Wu Y, Karlsson R, Barbalic M, Lin H, Pool R, Zhu G, Mace A, Sidore C, Trompet S, Mangino M, Sabater-Lleal M, Kemp JP, Abbasi A, Kacprowski T, Verweij N, Smith AV, Huang T, Marzi C, Feitosa MF, Lohman KK, Kleber ME, Milaneschi Y, Mueller C, Huq M, Vlachopoulou E, Lyytikainen LP, Oldmeadow C, Deelen J, Perola M, Zhao JH, Feenstra B, LifeLines Cohort S, Amini M, Group CIW, Lahti J, Schraut KE, Fornage M, Suktitipat B, Chen WM, Li X, Nutile T, Malerba G, Luan J, Bak T, Schork N, Del Greco MF, Thiering E, Mahajan A, Marioni RE, Mihailov E, Eriksson J, Ozel AB, Zhang W, Nethander M, Cheng YC, Aslibekyan S, Ang W, Gandin I, Yengo L, Portas L, Kooperberg C, Hofer E, Rajan KB, Schurmann C, den Hollander W, Ahluwalia TS, Zhao J, Draisma HHM, Ford I, Timpson N, Teumer A, Huang H, Wahl S, Liu Y, Huang J, Uh HW, Geller F, Joshi PK, Yanek LR, Trabetti E, Lehne B, Vozzi D, Verbanck M, Biino G, Saba Y, Meulenbelt I, O'Connell JR, Laakso M, Giulianini F, Magnusson PKE, Ballantyne CM, Hottenga JJ, Montgomery GW, Rivadineira F, Rueedi R, Steri M, Herzig KH, Stott DJ, Menni C, Franberg M, St Pourcain B, Felix SB, Pers TH, Bakker SJL, Kraft P, Peters A, Vaidya D, Delgado G, Smit JH, Grossmann V, Sinisalo J, Seppala I, Williams SR, Holliday EG, Moed M, Langenberg C, Raikkonen K, Ding J, Campbell H, Sale MM, Chen YI, James AL, Ruggiero D, Soranzo N, Hartman CA, Smith EN, Berenson GS, Fuchsberger C, Hernandez D, Tiesler CMT, Giedraitis V, Liewald D, Fischer K, Mellstrom D, Larsson A, Wang Y, Scott WR, Lorentzon M, Beilby J, Ryan KA, Pennell CE, Vuckovic D, Balkau B, Concas MP, Schmidt R, Mendes de Leon CF, Bottinger EP, Kloppenburg M, Paternoster L, Boehnke M, Musk AW, Willemsen G, Evans DM, Madden PAF, Kahonen M, Kutalik Z, Zoledziewska M, Karhunen V, Kritchevsky SB, Sattar N, Lachance G, Clarke R, Harris TB, Raitakari OT, Attia JR, van Heemst D, Kajantie E, Sorice R, Gambaro G, Scott RA, Hicks AA, Ferrucci L, Standl M, Lindgren CM, Starr JM, Karlsson M, Lind L, Li JZ, Chambers JC, Mori TA, de Geus E, Heath AC, Martin NG, Auvinen J, Buckley BM, de Craen AJM, Waldenberger M, Strauch K, Meitinger T, Scott RJ, McEvoy M, Beekman M, Bombieri C, Ridker PM, Mohlke KL, Pedersen NL, Morrison AC, Boomsma DI, Whitfield JB, Strachan DP, Hofman A, Vollenweider P, Cucca F, Jarvelin MR, Jukema JW, Spector TD, Hamsten A, Zeller T, Uitterlinden AG, Nauck M, Gudnason V, Qi L, Grallert H, Borecki IB, Rotter JI, Marz W, Wild PS, Lokki ML, Boyle M, Salomaa V, Melbye M, Eriksson JG, Wilson JF, Penninx B, Becker DM, Worrall BB, Gibson G, Krauss RM, Ciullo M, Zaza G, Wareham NJ, Oldehinkel AJ, Palmer LJ, Murray SS, Pramstaller PP, Bandinelli S, Heinrich J, Ingelsson E, Deary IJ, Magi R, Vandenput L, van der Harst P, Desch KC, Kooner JS, Ohlsson C, Hayward C, Lehtimaki T, Shuldiner AR, Arnett DK, Beilin LJ, Robino A, Froguel P, Pirastu M, Jess T, Koenig W, Loos RJF, Evans DA, Schmidt H, Smith GD, Slagboom PE, Eiriksdottir G, Morris AP, Psaty BM, Tracy RP, Nolte IM, Boerwinkle E, Visvikis-Siest S, Reiner AP, Gross M, Bis JC, Franke L, Franco OH, Benjamin EJ, Chasman DI, Dupuis J, Snieder H, Dehghan A and Alizadeh BZ. Genome Analyses of >200,000 Individuals Identify 58 Loci for Chronic Inflammation and Highlight Pathways that Link Inflammation and Complex Disorders. *Am J Hum Genet*. 2018;103:691-706.
2. de Vries PS, Chasman DI, Sabater-Lleal M, Chen MH, Huffman JE, Steri M, Tang W, Teumer A, Marioni RE, Grossmann V, Hottenga JJ, Trompet S, Muller-Nurasyid M, Zhao JH, Brody JA, Kleber ME, Guo X, Wang JJ, Auer PL, Attia JR, Yanek LR, Ahluwalia TS, Lahti J, Venturini C, Tanaka T, Bielak LF, Joshi PK, Rocanin-Arjo A, Kolcic I, Navarro P, Rose LM, Oldmeadow C, Riess H, Mazur J, Basu S, Goel A, Yang Q, Ghanbari M, Willemsen G, Rumley A, Fiorillo E, de Craen AJ, Grotevendt A, Scott R, Taylor KD, Delgado GE, Yao J, Kifley A, Kooperberg C, Qayyum R, Lopez LM, Berentzen TL, Raikkonen K, Mangino M, Bandinelli S, Peyser PA, Wild S, Tregouet DA, Wright AF, Marten J, Zemunik T, Morrison AC, Sennblad B, Tofler G, de Maat MP, de Geus EJ, Lowe GD, Zoledziewska M, Sattar N, Binder H, Volker U, Waldenberger M, Khaw KT, McKnight B, Huang J, Jenny NS, Holliday EG, Qi L, McEvoy MG,

Becker DM, Starr JM, Sarin AP, Hysi PG, Hernandez DG, Jhun MA, Campbell H, Hamsten A, Rivadeneira F, McArdle WL, Slagboom PE, Zeller T, Koenig W, Psaty BM, Haritunians T, Liu J, Palotie A, Uitterlinden AG, Stott DJ, Hofman A, Franco OH, Polasek O, Rudan I, Morange PE, Wilson JF, Kardia SL, Ferrucci L, Spector TD, Eriksson JG, Hansen T, Deary IJ, Becker LC, Scott RJ, Mitchell P, Marz W, Wareham NJ, Peters A, Greinacher A, Wild PS, Jukema JW, Boomsma DI, Hayward C, Cucca F, Tracy R, Watkins H, Reiner AP, Folsom AR, Ridker PM, O'Donnell CJ, Smith NL, Strachan DP and Dehghan A. A meta-analysis of 120 246 individuals identifies 18 new loci for fibrinogen concentration. *Hum Mol Genet*. 2016;25:358-70.

3. Ahola-Olli AV, Wurtz P, Havulinna AS, Aalto K, Pitkanen N, Lehtimaki T, Kahonen M, Lyytikainen LP, Raitoharju E, Seppala I, Sarin AP, Ripatti S, Palotie A, Perola M, Viikari JS, Jalkanen S, Maksimow M, Salomaa V, Salmi M, Kettunen J and Raitakari OT. Genome-wide Association Study Identifies 27 Loci Influencing Concentrations of Circulating Cytokines and Growth Factors. *Am J Hum Genet*. 2017;100:40-50.

4. Sun BB, Maranville JC, Peters JE, Stacey D, Staley JR, Blackshaw J, Burgess S, Jiang T, Paige E, Surendran P, Oliver-Williams C, Kamat MA, Prins BP, Wilcox SK, Zimmerman ES, Chi A, Bansal N, Spain SL, Wood AM, Morrell NW, Bradley JR, Janjic N, Roberts DJ, Ouwehand WH, Todd JA, Soranzo N, Suhre K, Paul DS, Fox CS, Plenge RM, Danesh J, Runz H and Butterworth AS. Genomic atlas of the human plasma proteome. *Nature*. 2018;558:73-79.

5. Shim H, Chasman DI, Smith JD, Mora S, Ridker PM, Nickerson DA, Krauss RM and Stephens M. A multivariate genome-wide association analysis of 10 LDL subfractions, and their response to statin treatment, in 1868 Caucasians. *PLoS One*. 2015;10:e0120758.

6. Astle WJ, Elding H, Jiang T, Allen D, Ruklisa D, Mann AL, Mead D, Bouman H, Riveros-Mckay F, Kostadima MA, Lambourne JJ, Sivapalaratnam S, Downes K, Kundu K, Bomba L, Berentsen K, Bradley JR, Daugherty LC, Delaneau O, Freson K, Garner SF, Grassi L, Guerrero J, Haimel M, Janssen-Megens EM, Kaan A, Kamat M, Kim B, Mandoli A, Marchini J, Martens JHA, Meacham S, Megy K, O'Connell J, Petersen R, Sharifi N, Sheard SM, Staley JR, Tuna S, van der Ent M, Walter K, Wang SY, Wheeler E, Wilder SP, Iotchkova V, Moore C, Sambrook J, Stunnenberg HG, Di Angelantonio E, Kaptoge S, Kuijpers TW, Carrillo-de-Santa-Pau E, Juan D, Rico D, Valencia A, Chen L, Ge B, Vasquez L, Kwan T, Garrido-Martin D, Watt S, Yang Y, Guigo R, Beck S, Paul DS, Pastinen T, Bujold D, Bourque G, Frontini M, Danesh J, Roberts DJ, Ouwehand WH, Butterworth AS and Soranzo N. The Allelic Landscape of Human Blood Cell Trait Variation and Links to Common Complex Disease. *Cell*. 2016;167:1415-1429 e19.

7. Scott RA, Lagou V, Welch RP, Wheeler E, Montasser ME, Luan J, Magi R, Strawbridge RJ, Rehnberg E, Gustafsson S, Kanoni S, Rasmussen-Torvik LJ, Yengo L, Lecoeur C, Shungin D, Sanna S, Sidore C, Johnson PC, Jukema JW, Johnson T, Mahajan A, Verweij N, Thorleifsson G, Hottenga JJ, Shah S, Smith AV, Sennblad B, Gieger C, Salo P, Perola M, Timpson NJ, Evans DM, Pourcain BS, Wu Y, Andrews JS, Hui J, Bielak LF, Zhao W, Horikoshi M, Navarro P, Isaacs A, O'Connell JR, Stirrups K, Vitart V, Hayward C, Esko T, Mihailov E, Fraser RM, Fall T, Voight BF, Raychaudhuri S, Chen H, Lindgren CM, Morris AP, Rayner NW, Robertson N, Rybin D, Liu CT, Beckmann JS, Willems SM, Chines PS, Jackson AU, Kang HM, Stringham HM, Song K, Tanaka T, Peden JF, Goel A, Hicks AA, An P, Muller-Nurasyid M, Franco-Cereceda A, Folkersen L, Marullo L, Jansen H, Oldehinkel AJ, Bruinenberg M, Pankow JS, North KE, Forouhi NG, Loos RJ, Edkins S, Varga TV, Hallmans G, Oksa H, Antonella M, Nagaraja R, Trompet S, Ford I, Bakker SJ, Kong A, Kumari M, Gigante B, Herder C, Munroe PB, Caulfield M, Antti J, Mangino M, Small K, Miljkovic I, Liu Y, Atalay M, Kiess W, James AL, Rivadeneira F, Uitterlinden AG, Palmer CN, Doney AS, Willemsen G, Smit JH, Campbell S, Polasek O, Bonnycastle LL, Hercberg S, Dimitriou M, Bolton JL, Fowkes GR, Kovacs P, Lindstrom J, Zemunik T, Bandinelli S, Wild SH, Basart HV, Rathmann W, Grallert H, Replication DIG, Meta-analysis C, Maerz W, Kleber ME, Boehm BO, Peters A, Pramstaller PP, Province MA, Borecki IB, Hastie ND, Rudan I, Campbell H, Watkins H, Farrall M, Stumvoll M, Ferrucci L, Waterworth DM, Bergman RN, Collins FS, Tuomilehto J, Watanabe RM, de Geus EJ, Penninx BW, Hofman

A, Oostra BA, Psaty BM, Vollenweider P, Wilson JF, Wright AF, Hovingh GK, Metspalu A, Uusitupa M, Magnusson PK, Kyvik KO, Kaprio J, Price JF, Dedoussis GV, Deloukas P, Meneton P, Lind L, Boehnke M, Shuldiner AR, van Duijn CM, Morris AD, Toenjes A, Peyser PA, Beilby JP, Korner A, Kuusisto J, Laakso M, Bornstein SR, Schwarz PE, Lakka TA, Rauramaa R, Adair LS, Smith GD, Spector TD, Illig T, de Faire U, Hamsten A, Gudnason V, Kivimaki M, Hingorani A, Keinanen-Kiukaanniemi SM, Saaristo TE, Boomsma DI, Stefansson K, van der Harst P, Dupuis J, Pedersen NL, Sattar N, Harris TB, Cucca F, Ripatti S, Salomaa V, Mohlke KL, Balkau B, Froguel P, Pouta A, Jarvelin MR, Wareham NJ, Bouatia-Naji N, McCarthy MI, Franks PW, Meigs JB, Teslovich TM, Florez JC, Langenberg C, Ingelsson E, Prokopenko I and Barroso I. Large-scale association analyses identify new loci influencing glycemic traits and provide insight into the underlying biological pathways. *Nat Genet*. 2012;44:991-1005.

8. Kettunen J, Demirkan A, Wurtz P, Draisma HH, Haller T, Rawal R, Vaarhorst A, Kangas AJ, Lyytikainen LP, Pirinen M, Pool R, Sarin AP, Soininen P, Tukiainen T, Wang Q, Tiainen M, Tynkkynen T, Amin N, Zeller T, Beekman M, Deelen J, van Dijk KW, Esko T, Hottenga JJ, van Leeuwen EM, Lehtimaki T, Mihailov E, Rose RJ, de Craen AJ, Gieger C, Kahonen M, Perola M, Blankenberg S, Savolainen MJ, Verhoeven A, Viikari J, Willemsen G, Boomsma DI, van Duijn CM, Eriksson J, Jula A, Jarvelin MR, Kaprio J, Metspalu A, Raitakari O, Salomaa V, Slagboom PE, Waldenberger M, Ripatti S and Ala-Korpela M. Genome-wide study for circulating metabolites identifies 62 loci and reveals novel systemic effects of LPA. *Nat Commun*. 2016;7:11122.
